# CRISPR-based assays for point of need detection and subtyping of influenza

**DOI:** 10.1101/2023.05.26.23290593

**Authors:** Yibin B. Zhang, Jon Arizti-Sanz, A’Doriann Bradley, Tinna-Solveig F. Kosoko-Thoroddsen, Pardis C. Sabeti, Cameron Myhrvold

## Abstract

The high disease burden of influenza virus poses a significant threat to human health and requires better methods to rapidly detect its many circulating species, subtypes, and variants. No current diagnostic technology meets the combined critical needs for a rapid, sensitive, specific, and cost-effective method for point-of-need (PON) influenza detection and discrimination with minimal equipment requirements. Here, we introduce such a method using SHINE (Streamlined Highlighting of Infections to Navigate Epidemics), a CRISPR-based RNA detection platform. We develop and validate four SHINE assays for the detection and differentiation of clinically relevant influenza species (A and B) and subtypes (H1N1 and H3N2). These optimized assays achieve 100% concordance with reverse-transcriptase real-time polymerase chain reaction (RT-qPCR) when tested on clinical samples. We also created duplex Cas12/Cas13 SHINE assays to simultaneously detect two targets and demonstrate its use in discriminating two alleles of an oseltamivir resistance (H275Y) mutation as well as to detect influenza A and human RNAse P, as a built-in internal control. Our assays have the potential to expand influenza detection outside of clinical laboratories in order to enhance influenza diagnosis and surveillance.

## Introduction

Influenza poses a major threat to public health, in terms of both its disease burden in seasonal outbreaks and its ability to cause pandemics. An estimated 1 billion people are infected annually with influenza A virus (IAV) subtypes H1N1 and H3N2 and influenza B virus (IBV), resulting in 3-5 million cases of severe disease and over 300,000 deaths globally [1]. The clinical presentation and disease severity of influenza depends on its species and subtype. For example, infections with H3N2 IAV are associated with more severe disease, including leukopenia and high fevers [2]. Occasionally, zoonotic influenza strains spill over into human populations with devastating consequences. Influenza pandemics in 1889 (H3N8), 1918 (H1N1), 1957 (H2N2), 1968 (H3N2), 1977 (H1N1) and 2009 (H1N1) each led to over 1 million deaths worldwide [3–5]. Real-time species and subtype information at the point of need (PON) can guide clinical decision-making and allow for the rapid implementation of infection prevention and control measures [6,7].

Viral sequence data is particularly important in the context of drug-resistant influenza strains [8]. Oseltamivir, a neuraminidase (NA) inhibitor, is one of the most widely used antivirals for seasonal influenza and is heavily stockpiled for pandemic response. However, a single nucleotide polymorphism (SNP) causing the H275Y amino acid substitution in the NA gene confers oseltamivir-resistance to H1N1 IAV in most clinically observed cases, and its prevalence is increasing worldwide [9,10]. Expanded detection of the many circulating species, subtypes, and variants of influenza virus is critical for effective clinical care, surveillance, and outbreak response.

Constraints of current diagnostic methods hinder their deployment and utility at the PON as diagnostic testing for influenza relies on laboratory infrastructure [11]. More than 80,000 patient samples are collected and tested for influenza annually in the United States. However, this figure pales in comparison to the estimated case load of 26 million, underscoring the need for more accessible point-of-need (PON) diagnostic technologies that can bridge the gap and improve testing and data collection for monitoring its spread [12]. The current gold standard for laboratory-based influenza virus detection, reverse-transcription quantitative polymerase chain reaction (RT-qPCR), is challenging to deploy at PON due to high costs, assay complexity and equipment requirements [13,14]. Isothermal methods, such as reverse-transcription loop-mediated isothermal amplification (RT-LAMP), are fast and have higher sensitivity than antigen-based tests, but they currently require auxiliary devices for sample processing and heating, limiting their utility at the PON [15]. Antigen-capture tests are quick and user-friendly, but their moderate sensitivity results in false negatives and underdiagnosis of potentially infectious individuals [16].

Similarly, characterization of subtypes and variants is currently limited. Sequencing is the gold standard method for detecting influenza mutations, but can cost upwards of hundreds of dollars per sample and take days to weeks to return results; and is therefore rarely performed in clinical laboratories [13,17]. In addition, RT-qPCR-based genotyping assays (e.g. for H275Y) are not widely available, even in high-resource laboratories and clinical settings [17–19]. Therefore, there is an unmet need for diagnostic technologies that are accurate, user-friendly, cost-effective, clinically relevant, and capable of providing genetic information about influenza virus.

Recently developed CRISPR-based diagnostic assays (CRISPR-Dx) present a promising approach for influenza virus detection and discrimination. CRISPR-Dx are highly specific, sensitive, and programmable, as they rely on complementary base pairing between a CRISPR RNA (crRNA) and the target of interest to activate the nuclease activity of the CRISPR-associated Cas12 or Cas13 nucleases [20–22]. Upon activation, the Cas12 or Cas13 complexes can cleave DNA- or RNA-based reporters, respectively, leading to a fluorescent or colorimetric, paper-based signal (Figure 1A). Given their excellent specificity, CRISPR-Dx are also uniquely suited to detect SNPs. SNP discrimination is achieved by measuring relative signal intensities between a set of two crRNAs, one specific to each allele [20,23–27]. In addition, the orthogonality of Cas12- and Cas13-based systems allows their integration into a single reaction, allowing simultaneous detection of two targets from a single sample [24,28,29].

**Figure 1.**
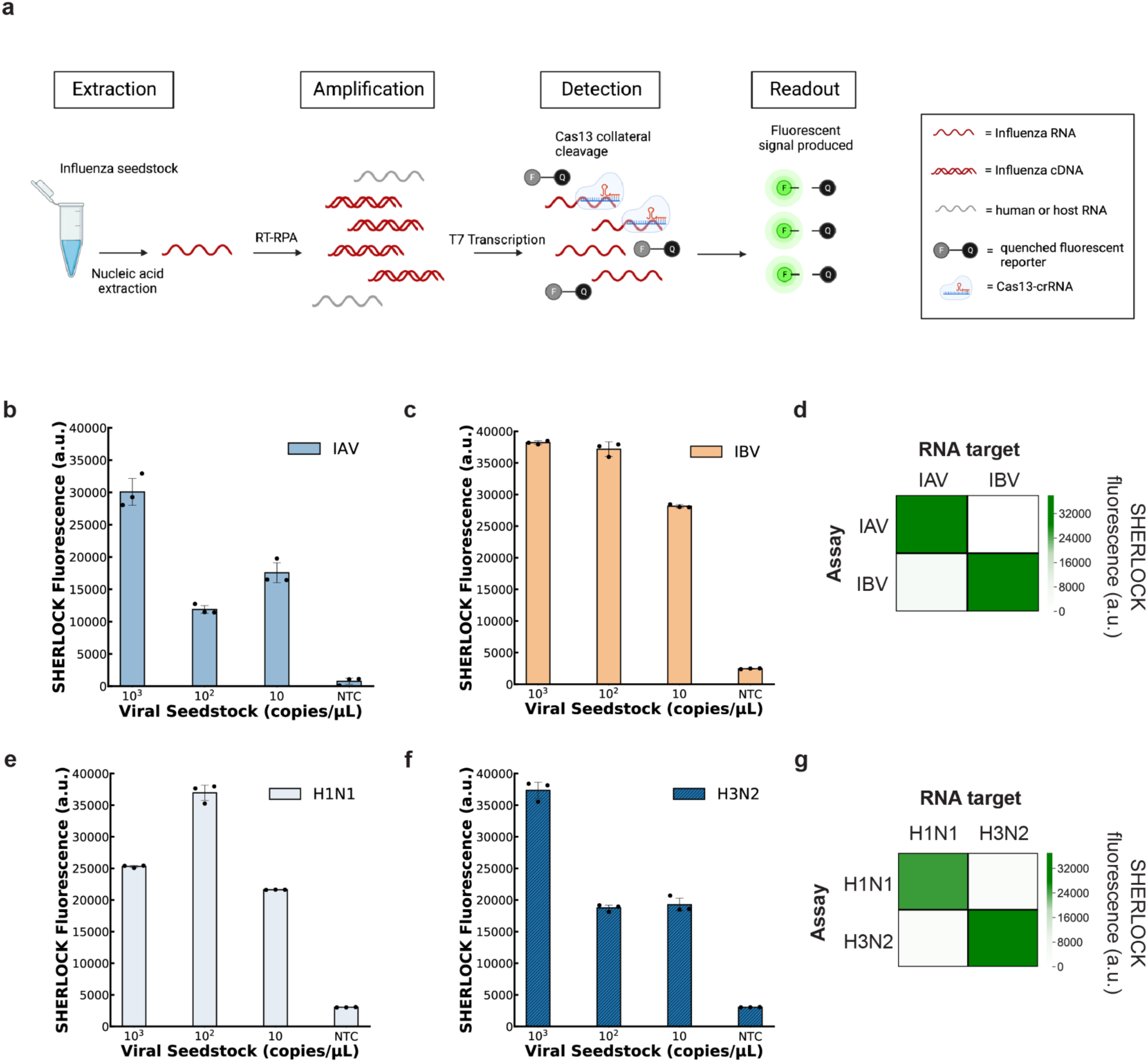
SHERLOCK assays for influenza detection and subtyping. **a)** Schematic of SHERLOCK workflow, including nucleic acid extraction, nucleic acid amplification with reverse-transcription recombinase polymerase amplification (RT-RPA), T7 transcription and CRISPR-Cas13-mediated cleavage of a quenched fluorescence reporter. **b,c)** SHERLOCK fluorescence of (**b**) IAV and (**c**) IBV assays on a serial dilution of viral seedstock RNA. **d)** Heatmap showing the specificity of the IAV and IBV SHERLOCK assays on target RNA at 10^5^ cp/µL. **e,f)** SHERLOCK fluorescence of (**e**) H1N1 and (**f**) H3N2 assays on a serial dilution of viral seedstock RNA. **g)** Heatmap showing the specificity of the H1N1 and H3N2 SHERLOCK assays on viral seedstock RNA at 10^5^ cp/µL. For **b,c,e** and **f**, values are mean fluorescence ± standard deviation of 3 technical replicates after 1.5 h. In **d** and **g**, heatmap values represent the mean of 3 technical replicates. NTC, no target control.

In recent years, technical advances in laboratory-based CRISPR-Dx have simplified their use and increased their utility at the PON [23–25,30–34]. SHINE [24] (Streamlined Highlighting of Infections to Navigate Epidemics), for instance, is an end-to-end diagnostic platform that builds upon the laboratory-based SHERLOCK (Specific High-sensitivity Enzymatic Reporter unLOCKing) technology by incorporating fast and ambient temperature sample processing as well as freeze-dried detection reagents for increased usability and deployability. We previously demonstrated SHINE is a simple, accurate and highly modular technology with the capacity to detect SARS-CoV-2 and identify clinically-relevant mutations, with minimal equipment requirements [24,30]. As such, this platform has the potential to become a widely distributed PON diagnostic test for seasonal and pandemic influenza strains.

Here, we present the development of CRISPR-Dx assays to detect and differentiate between clinically relevant species and subtypes of influenza, namely, IAV and IBV, as well as IAV subtypes H1N1 and H3N2 [35]. We build upon the recently published SHINE technology and show that influenza virus detection can be achieved with minimal equipment requirements using a paper-based readout, facilitating its PON deployment [24]. We also describe a method for multiplexing Cas13- and Cas12-based assays on the simplified, single-reaction SHINE platform and developed a highly accurate duplex assay that can identify the presence of oseltamivir-resistance in clinical samples. Further leveraging this platform, we also developed a dual influenza A and Human RNase P duplex detection assay; the latter can serve as an internal control in clinical testing. This set of influenza assays presents a promising avenue toward rapid, low-cost, and straightforward diagnostics that can enable the widespread detection and surveillance of influenza.

## Results

### Design and testing of laboratory-based SHERLOCK influenza assays

We sought to develop clinically relevant CRISPR-Dx assays for influenza virus, focusing on the detection of IAV and IBV as well as the discrimination of IAV subtypes H1N1 and H3N2. To design the sequences of crRNAs and primers for our assays, we used ADAPT (Activity-informed Design with All-inclusive Patrolling of Targets), a software platform that leverages machine learning algorithms to maximize sensitivity and target specificity in Cas13-based assay design [36]. We identified sets of crRNAs and primer pairs with high predicted activity *in silico* located in segments 2, 4 and 7 of the influenza genome that encode for the polymerase basic 1 (PB1), matrix protein (MP) and hemagglutinin (HA) proteins, respectively. We selected the three highest scoring primer-crRNA sets for each of the four assays for further testing.

We validated our CRISPR-Dx assays for SHERLOCK using synthetic RNA targets and viral seedstocks. We first tested the 3 highest predicted activity primer-crRNA sets designed for each of our 4 assays (IAV, IBV, H1N1, H3N2) against matching synthetic RNA targets for each. We selected the best performing primer-crRNA set for each assay based on the limit of detection (LoD) and reaction kinetics, and these are henceforth referred to as the IAV, IBV, H1N1 or H3N2 assays (Supplementary Figures 1 and 2). Each of these assays was able to detect synthetic influenza RNA targets down to a concentration of 10 copies/µL within 1.5 hours. To further validate these results, we tested the IAV and IBV SHERLOCK assays against a serial dilution of RNA extracted from IAV (H1N1) and IBV viral seedstocks respectively, demonstrating sensitive detection down to 10 copies/µL (Figure 1B, C). Similarly, the H1N1 and H3N2 SHERLOCK assays detected extracted viral RNA (Figure 1E, F) down to a concentration of 10 copies/µL within 1.5 hours.

The four SHERLOCK influenza assays comprehensively capture the species and subtype sequence diversity of IAV, IBV, H1N1, and H3N2, while retaining excellent specificity to their targeted species or subtype. Based on publicly available genomic information, the crRNA sequence of the IAV SHERLOCK assay was fully conserved in 99.8% of 15,688 IAV genomes analyzed, while the primers designed by ADAPT were predicted to amplify 99% of these genomes. Similarly, the crRNAs were predicted to target 99.8%, 99.7% and 99.7% of the analyzed 6422 IBV, 5509 H1N1 and 9209 H3N2 genomes, respectively; and ADAPT primers for each clade were predicted to amplify 99.5%, 99.8% and 99.6% of the genomes, respectively. Experimentally, each assay demonstrated excellent specificity, with high fluorescence on the desired target and negligible fluorescence on non-preferred targets, even at high (10^5^ copies/µL) RNA target concentrations (Figure 1D, G). These results demonstrate the development of highly sensitive and specific SHERLOCK assays for the detection of influenza species and subtypes suitable for use in laboratory settings.

### Development of PON influenza assays using SHINE

We next adapted our laboratory-based SHERLOCK influenza assays into the more user-friendly and minimally equipped SHINE platform to allow for a PON simple, streamlined workflow. We incorporated the amplification and detection steps of SHERLOCK into a single reaction, as previously described [30]. In this format, the resulting single-reaction assays were able to detect synthetic RNA from IAV and IBV; as well as IAV subtypes H1N1 and H3N2 down to a concentration of 100 copies/µL, using a fluorescence readout (Supplementary Figure 3).

We further simplified our influenza assays by freeze drying the reagents according to published protocol and validating performance [24]. The resulting lyophilized pellets can be rehydrated as needed, facilitating assay transportation and storage, increasing the ease-of-use, and reducing hands-on preparation time. The resulting SHINE assays were able to detect their intended synthetic RNA targets with favorable kinetics and high sensitivity using both fluorescence and paper-based readouts down to 100 copies/µL (Supplementary Figures 4-6). The assays demonstrated high specificity with high fluorescence against the desired target and negligible fluorescence against non-preferred targets, even at high (10^5^ copies/µL) viral RNA concentrations (Figure 2B). Against viral seedstocks, the lyophilized assays detected and differentiated H1N1, H3N2 and influenza B down to 100 copies/µL with both fluorescence and paper-based readouts (Figure 2C, D). This sensitivity is within the range required to detect influenza in most clinical samples based on reported patient NP swab viral loads, which are typically between 100 to 100,000 copies/µL, with a median of 10,000 copies/µL [37].

**Figure 2.**
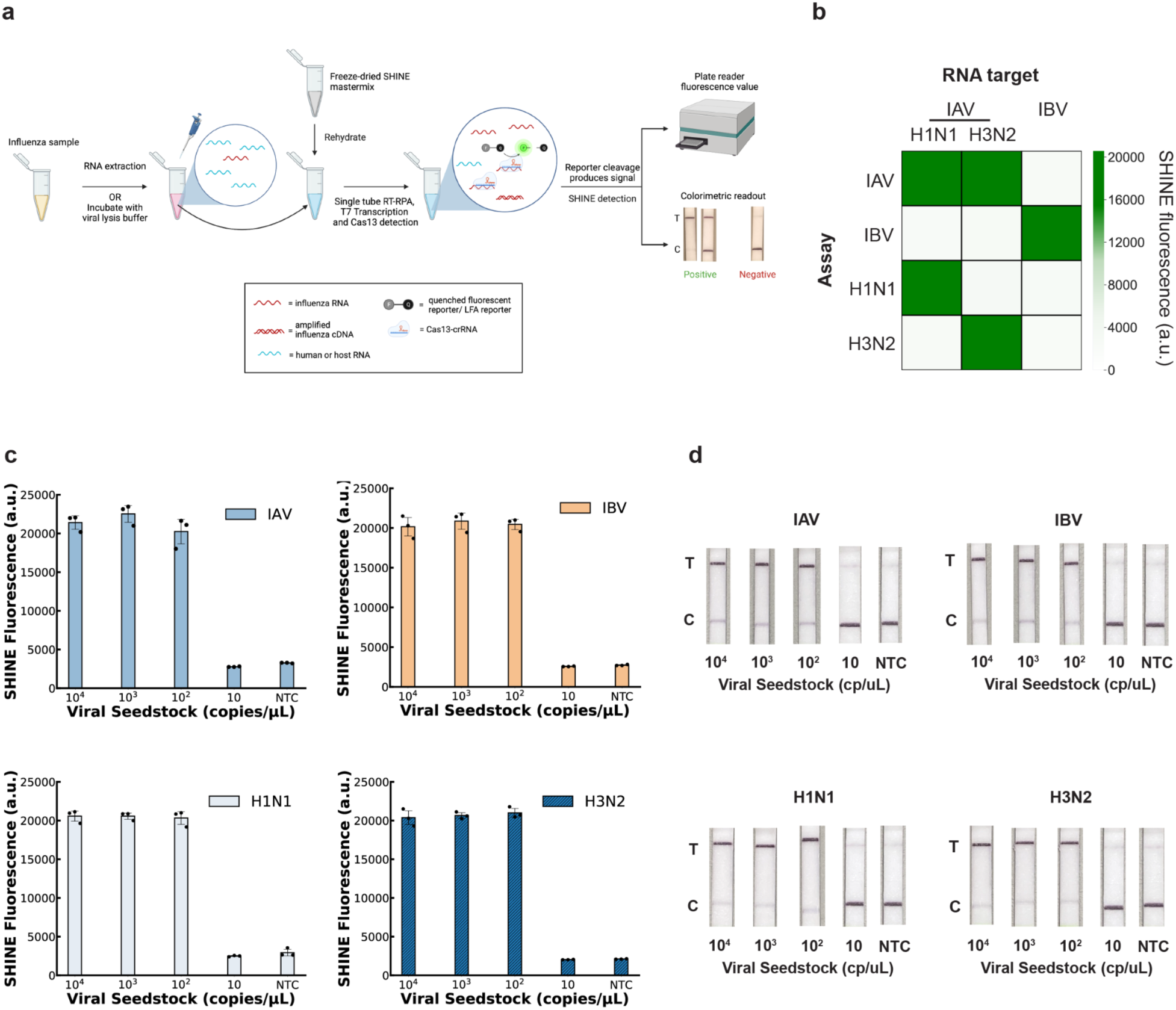
SHINE assays for influenza detection and subtyping. **a)** Schematic of SHINE workflow, including RNA extraction or ambient temperature inactivation of unextracted samples, and single-step amplification and detection with either a fluorescence-based or colorimetric, paper-based readout. For paper-based readouts, samples that contain the target should display a top test (T) band, which may be accompanied by a bottom (C) band if there is lower target concentration within the sample. Samples negative for the tested target should display a strong control (C) band and no significant top test (T) band. RT-RPA, reverse transcription recombinase polymerase amplification. **b)** Heatmap showing the specificity of the IAV, IBV, H1N1 and H3N2 SHINE assays on RNA extracted from viral seedstocks. Target RNA concentration: 10^5^ cp/µL. Fluorescence values represent the mean of 3 technical replicates after 1.5 h. **c)** Fluorescence readout of IAV, IBV, H1N1 and H3N2 SHINE assays on a serial dilution of RNA extracted from viral seedstocks. Values are mean fluorescence ± standard deviation of 3 technical replicates after 1.5 h. **c)** Paper-based colorimetric readout of IAV, IBV, H1N1 and H3N2 SHINE assays on a serial dilution of RNA extracted from viral seedstocks after 1.5 h incubation. Shown are representative images from 3 technical replicates. NTC, no target control.

Given the excellent performance of our SHINE assays on contrived samples, we sought to assess their clinical performance on a set of 25 extracted nasopharyngeal (NP) swab samples, using the US Center for Disease Control and Prevention (CDC)-recommended RT-qPCR protocols as the gold-standard, comparator tests [38]. We conducted side-by-side testing on this set of 5 IAV-positive samples (2 H1N1, 3H3N2), 10 IBV-positive samples and 10 influenza-negative samples. Our SHINE assays showed perfect (100%) concordance with RT-qPCR on all the samples tested, with both the fluorescence and paper-based readouts (Figure 3A, B; Supplementary Figure 7 and 8). In particular, our IAV and IBV SHINE assays were able to detect and differentiate the species of IAV and IBV positive samples, demonstrating 100% sensitivity and specificity. Of the IAV-positive samples, our H1N1 and H3N2 SHINE assays were also able to detect and differentiate between samples positive for these two key subtypes, demonstrating 100% sensitivity and specificity. All samples negative for influenza virus by RT-qPCR were also negative by the SHINE assays. These results show that even with simple, PON capable sample processing, the assays retained good performance, demonstrating its efficacy as a streamlined influenza detection platform with minimal equipment requirements.

**Figure 3.**
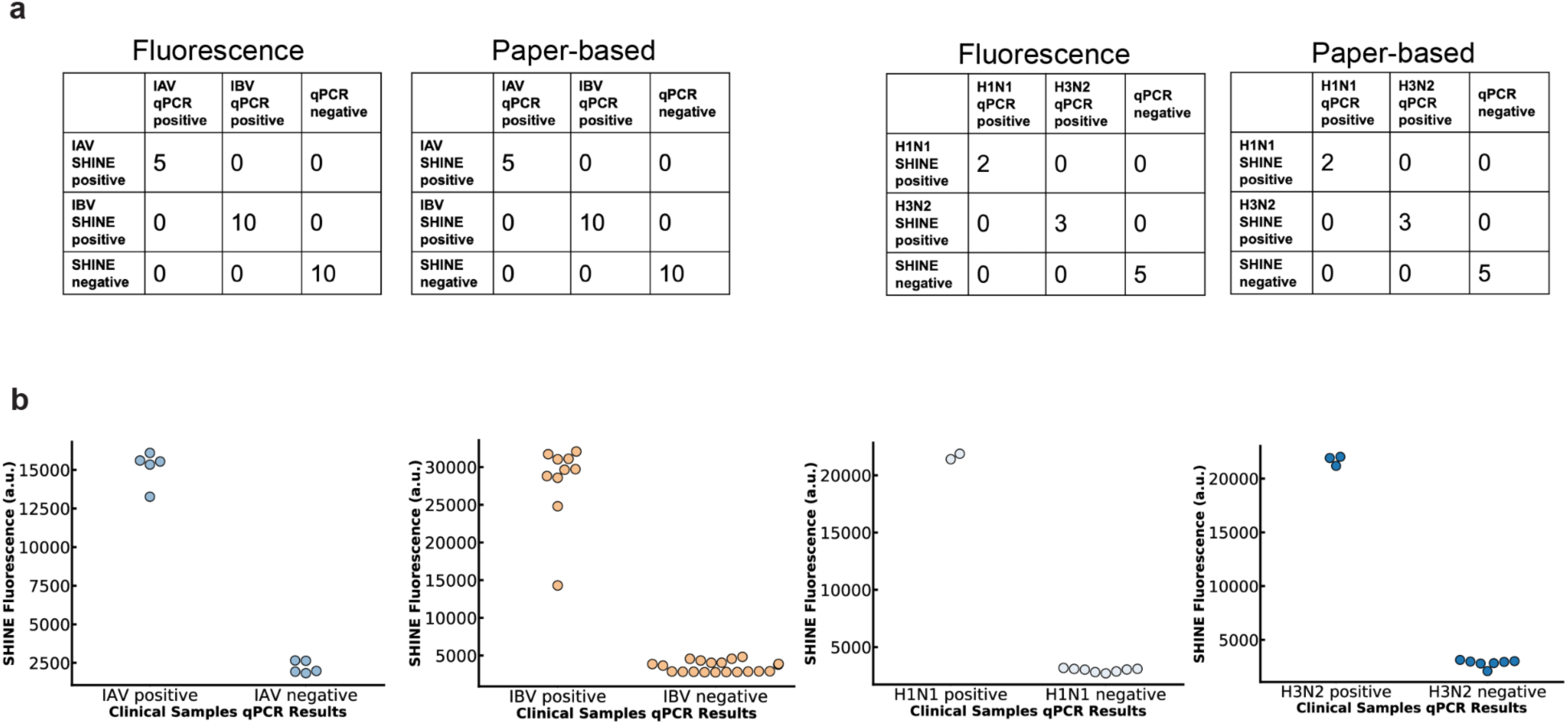
Evaluation of SHINE influenza assay performance against clinical samples. **a)** Summary of concordance between SHINE and RT-qPCR detection in both fluorescence and paper-based readouts in viral detection from patient NP swabs. All SHINE assays were 100% concordant with RT-qPCR results. **b)** Mean fluorescence values of SHINE assays detecting IAV, IBV, H1N1 and H3N2 from clinical samples. Values are mean fluorescence ± standard deviation of 3 technical replicates after 1.5 h.

### SHINE duplex influenza oseltamivir resistance assay

Given the importance of IAV drug resistance on clinical decision-making, we sought to design a duplex SHINE assay to simultaneously detect IAV and discriminate drug resistance SNP alleles. Initially, SNP detection with CRISPR-Dx was achieved using a single enzyme (e.g. Cas13) in two separate reactions, each containing a crRNA designed to detect either the wild-type or the mutated target [20,23]. Enabling SNP discrimination in a single reaction format further reduces the complexity of the assay and increases its clinical utility. The recent duplex SHINE platform achieves this simultaneous detection of two targets using the orthogonal DNA-based Cas12a reporter system and RNA-based CRISPR-Cas13a reporter system [24,28].

We designed SHINE assays to detect each allele of the SNP causing the H275Y amino acid substitution driving oseltamivir resistant H1N1 IAV. We designed two different crRNAs - one for Cas13 and one for Cas12 - specific against the ancestral oseltamivir-susceptible H275 and derived oseltamivir-resistant H275Y alleles, respectively and implemented it into two separate SHINE assays. We used a paper-based readout to assess assay performance on oseltamivir-susceptible H275 and oseltamivir-resistant H275Y seedstocks. The assays showed higher on-target than off-target activity for their respective allele, demonstrating the feasibility of SHINE in detecting oseltamivir-resistant IAV with minimal equipment requirements (Figure 4A).

**Figure 4:**
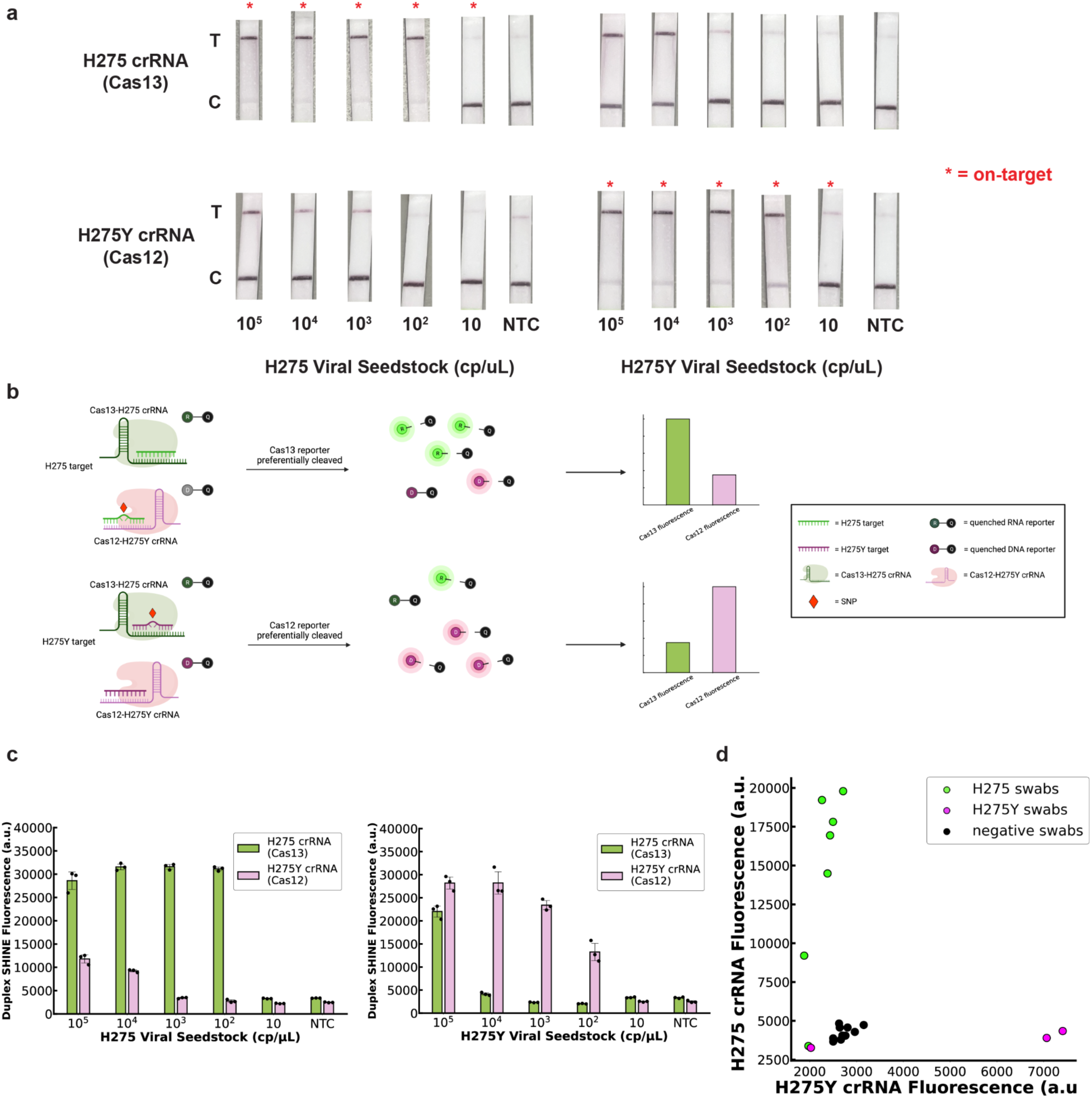
Development and characterization of a duplex H275Y oseltamivir-resistance assay. **a)** Paper-based readout of single-plex SHINE of the respective Cas13 crRNA targeting ancestral oseltamivir-susceptible H275 and Cas12 crRNA targeting derived oseltamivir-resistant H275Y alleles on H275 and H275Y seedstocks. **b)** Schematic of single nucleotide polymorphism (SNP) detection with CRISPR-Cas12 and Cas13. **c)** Detection of RNA extracted from H275 and H275Y viral seedstock using the duplex SHINE assay. **d)** Cas12 vs Cas13 fluorescence of extracted clinical NP swabs. For **c**, values are mean ± standard deviation for 3 technical replicates. For **d,** each point represents the mean Cas12 and Cas13 fluorescence of three technical replicates for a patient sample. All fluorescence measured after 1.5 h. NTC, no target control.

Having demonstrated that H275Y SNP detection was possible, we aimed to design H275Y SNP assays that can be run in a single reaction (Figure 4B). We designed two configurations: a duplexed assay in which Cas13 targets the H275 allele and Cas12 targets the H275Y allele and another assay in which Cas13 targets the H275Y allele and Cas12 targets the H275 allele (Supplementary Figure 9A, C). We tested these two configurations using synthetic RNA targets. We found the Cas13-H275 and Cas12-H275Y duplex SHINE assay to have increased specificity and more favorable detection kinetics; and therefore, we selected these assays for further development (Supplementary Figure 9B, D).

We lyophilized the duplex SHINE reaction and tested it on extracted RNA from oseltamivir-susceptible H275 and oseltamivir-resistant H275Y seedstocks. We detected RNA down to 100 copies/µL with good discrimination for both alleles (Figure 4C). Importantly, we observed that the fluorescence levels for Cas12 and Cas13 were higher for their respective, preferred targets than the non-preferred targets, especially in the 10^2^ to 10^4^ copies/µL range that is typical of influenza viral titers [37]. This would allow us to distinguish between H275 and H275Y H1N1 based on the ratio of fluorescence values obtained in the Cas12 and Cas13 channels.

We assessed the clinical performance of this IAV H275Y duplex SHINE assay. We compared performance against the gold-standard RT-qPCR assay on 20 patient NP swab samples with either H275 or H275Y H1N1, as confirmed by next generation sequencing (Figure 4D). Our duplex SHINE assay detected IAV RNA in 6 of 7 H275 and 2 of 3 H275Y IAV-positive samples and in none of the IAV-negative samples. The two H1N1-positive swabs (one H275 and one H275Y) that were negative by duplex SHINE had high RT-qPCR cycle values (Ct > 35), indicating these samples had low levels of IAV RNA, quantities below SHINE’s LoD and outside the normal range of 10^2^ to 10^4^ copies/µL for an influenza-positive sample. The IAV strain determined by our duplex SHINE assay on positive samples was 100% concordant with sequencing results. These results demonstrate the capability for our duplex SHINE assay to rapidly and accurately detect H275Y oseltamivir resistant IAV in clinical samples, overcoming a critical need in influenza diagnostics.

### Development of a duplex influenza A + Human RNase P SHINE assay

Given the effectiveness of the influenza assays we had developed, we aimed to leverage the duplex SHINE platform to bring these assays closer to clinical use by incorporating an internal control. Internal controls are a common requirement in the regulatory approval process of diagnostic tests, and are used to ensure adequate nucleic acid extraction and assay integrity [39,40]. Human RNase P is a routinely used internal control for influenza and other viral qPCR assays. Therefore, we sought to integrate a previously developed Cas12-based SHINE assay for RNase P with the Cas13-based IAV SHINE assay presented above, to serve as an internal positive control [24]. To do so, we optimized the SHINE reaction to boost the sensitivity in the duplex format, by reducing the concentration of ribonucleotide triphosphate (rNTPs), increasing the Cas12 reporter concentration, and extending the primer length for the RNase P assay.

We tested the performance of our duplex SHINE assay for IAV and an internal positive control, in both synthetic and patient samples. The assay detected extracted IAV RNA down to 100 copies/µL and synthetic RNase P targets down to 1,000 copies/µL (Figure 5B). This sensitivity is sufficient to detect human RNase P in most clinical NP swabs and saliva samples, as they typically have median concentrations of 1300 and 10,000 copies/µL of RNase P, respectively [41,42]. We assessed the performance of the duplex SHINE assay on a set of 10 NP swab samples, 5 positive and 5 negative for IAV. The assay detected IAV in 5/5 IAV-positive samples and 0/5 of the influenza-negative samples, demonstrating 100% concordance with RT-qPCR (Figure 5C). Furthermore, duplex SHINE detected RNase P in all 10 samples, showcasing its utility as an internal positive control.

**Figure 5:**
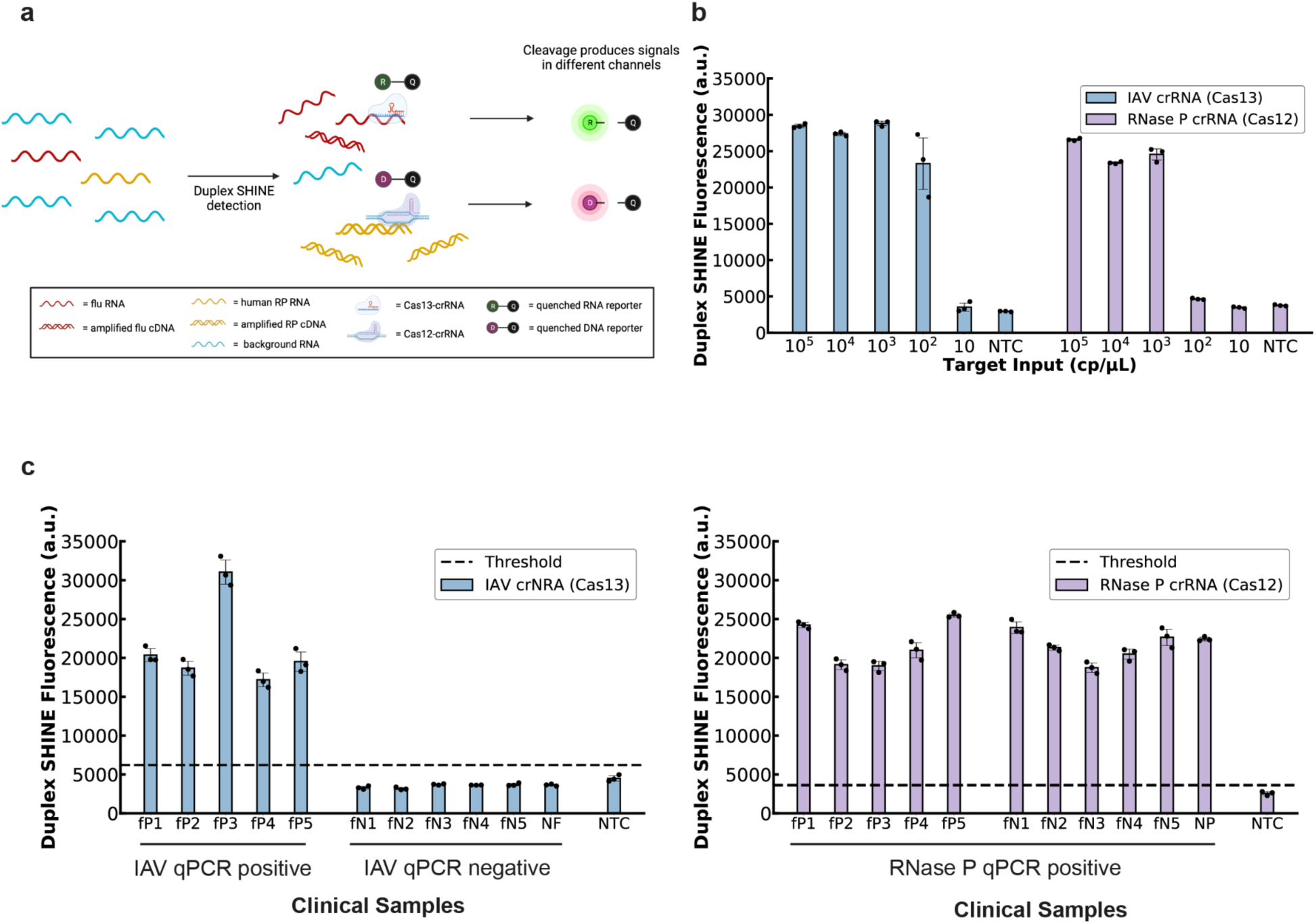
Development and characterization of duplex IAV + RNase P SHINE assay. **a)** Schematic of dual target detection in a single reaction using the orthogonal Cas13 and Cas12a enzymes. **b)** Detection of synthetic IAV and RNase P RNA targets with the duplex SHINE assay. **c)** Detection of nucleic acids extracted from nasopharyngeal (NP) swab samples and pooled human healthy nasal fluid with the duplex SHINE assay. NTC, no target control (i.e. universal transport medium used as input). NF, commercially purchased healthy pooled human nasal fluid. For **b** and **c**, values are mean fluorescence ± standard deviation for 3 technical replicates after 1.5h.

## Discussion

We leveraged recent developments in CRISPR-Dx to create highly specific and PON-deployable SHINE assays for the detection and differentiation of IAV, IBV, H1N1, H3N2, and oseltamivir-resistant strains, as well as the integration of an internal control. SHINE’s workflow is user-friendly, currently enabling the processing of 20 patient samples in under 20 minutes of hands-on time with results delivered in under 2 hours. As such, SHINE is well poised to expand viral detection and drug-resistance identification for influenza virus, especially in community clinics and resource-limited settings.

With the release of baloxavir to supplement existing oseltamivir influenza antivirals, there is now a clinical decision point on treatment options for influenza patients. Our SHINE assays have the potential to provide rapid detection of oseltamivir resistance, helping clinicians make better treatment choices at the PON.

SHINE is highly specific and programmable, allowing existing influenza assays to be easily adapted to new viral strains and variants and to new settings. Software platforms like ADAPT greatly simplify and automate the development of new SHINE assays to target new viral strains and variants as soon as viral genomes become publicly available [36]. SHINE assays could easily be expanded for the surveillance of influenza virus in livestock or be deployed in wastewater viral sampling - areas with strong economic incentives and important implications for human health [43,44].

Widely available diagnostic testing for influenza could enable better surveillance and inform antiviral drug prescription decisions, but additional advances will be required for CRISPR-Dx to be performed in virtually any location. Rapid (< 30 mins) and ambient temperature diagnostics with a visual readout and no equipment requirements would be ideal for deployment in remote and low-resourced environments. Solution-based, colorimetric approaches as well as amplification-free technologies could alleviate some of the common equipment requirements for CRISPR-Dx [45]. Similarly, the speed and sensitivity of CRISPR-Dx could be boosted using novel signal amplification schemes or with the use of auxiliary proteins [31]. With our SHINE influenza assays, we have taken steps to increase the accessibility and availability of influenza diagnostics, enabling better outbreak management and clinical treatment decisions.

## Methods

### Clinical samples and ethics statement

De-identified nasopharyngeal (NP) swabs (in UTM or VTM) from patients with influenza-like symptoms over the 2020-2021 influenza season (October - February) were provided to the Broad Institute by the CDC (USA). An additional non-human subjects research determination (NHSR-4318) and exempt determination (EX-7209) were made by the Broad Institute. This study was also reviewed by the CDC and was considered to not be human subject research. See Supplementary table 1 for additional clinical sample information.

### Design of Cas13 influenza assays

Using the National Center for Biotechnology Information (NCBI) Influenza database, publicly available human influenza sequences were downloaded in May 2019. Queries were limited to samples collected from May 2014 to May 2019 to ensure that the sequences are up to date and relevant for currently circulating strains. PB1 and MP segments of the viral genome were chosen for IAV and IBV testing, since these regions tend to be more conserved, allowing the designed assays to remain functional for a long time (<u>Mutational analysis of the conserved motifs of influenza A virus polymerase basic protein 1</u>; https://journals.plos.org/plosone/article?id=10.1371/journal.pone.0244669). These segments are also targeted by RT-qPCR assays designed by the World Health Organization (World Health Organization, 2020). H1N1 and H3N2 assays were designed to target the HA segment.

RPA primers and crRNA guides were designed for each of the four assays using ADAPT at commit version #f202977, a machine learning based guide design software [36]. The parameters were set as such: crRNA (oligo-length 28, mismatches 1, coverage-fraction 0.95, id-m 4) and RPA primer (oligo-length 30, mismatches 2, coverage-fraction 0.95, id-m 4). The definition of these parameters are as follows: “oligo-length” refers to the length of the crRNA or RPA primer. “mismatches” mean the maximum number of tolerated mismatches between the oligonucleotide probes (oligos) and the target sequences. “coverage-fraction” sets minimum fraction of sequence diversity within the viral alignment that the oligos must capture according to the other user-defined criterion. “id-m” is one less than the minimum number of mismatches of the oligos with regards to an outgroup target in the cases of differential subtyping (in this case IAV vs IBV and H1N1 vs H3N2), ensuring that the oligos are type-specific. ADAPT outputs all the crRNA-primer pairs that fulfill the input parameters and provides a predicted efficiency of the assay design on a scale of 0-1, with 1 being the highest. The top three scoring crRNA-primer pairs were chosen for each of the four IAV, IBV, H1N1 and H3N2 assays as candidates for further experimental evaluation – all candidate designs had a score greater than 0.9.

RPA primers and crRNAs for the oseltamivir resistance duplex assay were designed with a standard heuristic for Cas13- and Cas12-based detection: spacers were designed with the consensus sequences of all NCBI database ancestral and H275Y H1N1 NA segment sequences with the SNP in the space sequence (Chen et al., 2018; Metsky et al., 2022). Primers were designed from consensus sequences flanking the spacer.

### Nucleic acids, materials, and reagents

All RPA primers were ordered from Integrated DNA Technologies (IDT) as custom DNA oligos. Forward primers contained a T7 promoter sequence on the 5’ end to allow for transcription of the amplified DNA product into RNA that Cas13 can detect. The crRNAs were ordered as synthetic Alt-R CRISPR-RNA sequences from IDT. gBlock DNA gene fragments (IDT) with a T7 promoter were ordered as synthetic testing targets. gBlocks were *in vitro* transcribed using HiScribe™ T7 High Yield RNA Synthesis Kits (NEB) to create synthetic RNA targets, according to manufacturer instructions.

Healthy human nasal fluid was purchased from Lee Biosolutions and used as a control in the duplex assays. Influenza AH1N1pdm, AH3N2 and B quantitative viral seedstock were ordered from Zeptometrix. Oseltamivir resistant viral seedstock was also purchased from Zeptometrix as Influenza A H1N1pdm Virus Oseltamivir-R (Isolate 2) Culture Fluid (Heat Inactivated). The manufacturer has confirmed the H1N1pdm viral stock for the absence and the H1N1pdm Virus Oseltamivir-R viral stock for the presence of the H275Y cytosine to uracil mutation by sequencing. Contrived patient samples were created in-house by mixing viral seedstocks with universal transport medium (BD) at a 1:10 ratio.

### Two-step influenza detection assay

This protocol involved the amplification of nucleic acids with RT-RPA first, followed by the detection of the amplified material with Cas13. RPA reactions were carried out using commercially available TwistAmp Basic RPA kits (TwistDx) according to manufacturer specifications; except, 2 units/µL of SuperScript IV reverse transcriptase (Thermo Fisher) and 2 units/µL of Murine RNase Inhibitor (New England Biolabs, NEB) were added to the reaction. Forward and reverse RPA primers were used at a final concentration of 400 nM and magnesium acetate at 14 mM, as previously described [20]. Samples were mixed 1:9 with RT-RPA master mix and heated for 20 minutes at 41°C. RPA no input refers to using nuclease free water as sample input in the RPA reaction.

Cas13 detection reactions were performed based on published methods (Myhrvold et al., 2018). Briefly, a detection master mix was created with the following reagents: 45 nM LwaCas13a protein, 22.5 nM crRNA, 125 nM RNaseAlert v2 (Thermo Fisher Scientific), 1X cleavage buffer (CB; 80 mM Tris pH 7.5 and 2 mM DTT), 2 units/µL murine RNase inhibitor (NEB), 1.5 units/µL NextGen T7 RNA polymerase (Lucigen), 1 mM of each rNTP (NEB), and 9 mM MgCl_2_ (added last). The RT-RPA products were added in a 1:19 ratio to the detection master mix. The fluorescence intensity of each reaction was measured on a Cytation 5 plate reader (Biotek) using a monochromator with excitation at 485 nm and emission at 520 nm. Readings were recorded every 5 minutes for 1.5 hours at 37°C.

### One-step influenza detection assay

This protocol involved the amplification and detection of nucleic acids in a single reaction. A SHINE master mix was made consisting of 1X SHINE buffer (20 mM HEPES pH 8.0 with 60 mM KCl and 5% PEG-8000), 45 nM LwaCas13a protein, 62.5 nM 6rU FAM quenched reporter (for fluorescence readout) or 1µM 14rU FAM-Biotin reporter (for paper-based readout), 1 unit/µL murine RNase inhibitor, 2 mM of each rNTP, 1 unit/µL NextGen T7 RNA polymerase, and 120 nM forward and reverse RPA primers. This master mix was used to resuspend the TwistAmp Basic RPA pellets (1 pellet per 102 µL master mix). After RPA pellet resuspension, 2 units/µL SuperScript IV reverse transcriptase, 0.1 unit/µL RNase H (NEB) and 22.5 nM crRNA were added to the mastermix. Finally, magnesium acetate was added to reach a concentration of 14 mM to create the final master mix.

Samples were added in 1:19 with the final master mix. For the fluorescence-based readout, the fluorescence of each reaction was measured on a Cytation 5 plate reader (Biotek) using a monochromator with excitation at 485 nm and emission at 520 nm. Readings were recorded every 5 minutes for 1.5 hours at 37°C. For the paper-based readout, the reactions were incubated at 37°C for 1.5 hours, and then diluted 1:4 in HybriDetect Assay Buffer (Milenia Biotec). The buffered solution was incubated at room temperature for 5 minutes; after which, a HybriDetect1 lateral flow paper strip was added. Test images were taken 5 minutes after the addition of the strip using a smartphone camera. Lateral flow reactions that are positive for the template show a top test (T) fluorescence band on the paper strip after incubation and they may or may not show a bottom control (C) band [23]. Lateral flow reactions that are negative for the template only show a bottom (C) band. Lateral flow reactions are not designed to be quantitative.

### Lyophilized influenza SHINE assays

Lyophilized, single-step SHINEv.2 assays for influenza were prepared as previously described with some modifications [24]. An optimized protocol was developed for the lyophilization of our influenza SHINE assays and duplex SHINE assays to retain their maximum sensitivities. Briefly, 1X SHINE buffer was replaced with 1X LYO buffer (20 mM HEPES pH 8.0, 5% weight/volume sucrose and 150mM mannitol) and magnesium acetate was not added. The master mix was aliquoted into single reaction sizes (65.5µL), flash frozen by immersing in liquid nitrogen for 30 second and lyophilized overnight in a Labconco FreeZone 4.5L Benchtop Freeze Dryer under vacuum and at -50°C.

Prior to use, influenza SHINE pellets were resuspended with rehydration buffer (60 mM KCl, 3.5% PEG-8000 and 14 mM magnesium acetate). After resuspension, samples (synthetic RNA, contrived samples or clinical samples) were added in a 1:19 ratio to rehydrated SHINE pellets. Fluorescence detection or lateral flow based detection were performed as described above.

### Clinical sample testing with SHINE and RT-qPCR

The clinical performance of our influenza SHINE assays was assessed against the gold-standard RT-qPCR assays on a set of extracted nasopharyngeal swab samples. See supplementary table 1 for additional sample information. RNA was extracted from clinical samples using the QIAamp viral RNA mini kit (Qiagen) following the manufacturer’s instructions, using a starting volume of 100 μl per sample and eluted into 100 μl of nuclease-free water. Extracted RNA was used immediately or stored at −80 °C. RT-qPCR and SHINE testing were performed side-by-side.

RT-qPCR testing on extracted patient samples was performed using two CDC RT-qPCR kits – #FluIVD03-1 for IAV and IBV and #FluRUO-15 for H1N1 and H3N2 – according to CDC protocols and with a sample input volume of 3.5 µL [38]. These kits also included RT-qPCR reagents for RNase P as an internal control for proper extraction and to ensure no significant degradation has occurred. RT-qPCR reactions were incubated on a QuantStudio 6 (Applied Biosystems) with the following cycling conditions: hold at 25°C for 2min, reverse transcription at 50 °C for 15 min, polymerase activation at 95 °C for 2 min and 40 cycles with a denaturing step at 95 °C for 3 s followed by annealing and elongation steps at 60 °C for 30 s. Data were analyzed using the Standard Curve module of the Applied Biosystems analysis software. For each assay tested, any sample with a Ct value below 38 was considered positive.

Lyophilized SHINE pellets for IAV, IBV, H1N1 and H3N2 were prepared the day before testing, following the protocols outlined above. Pellets for both fluorescence and paper-based readouts were prepared separately. RNA extracted from patient samples was used as input into the rehydrated SHINE reaction mixes as described above. For the fluorescence-based readout, a sample was considered positive when its recorded fluorescence intensity value was higher than the mean plus 5 standard deviations of a negative control for that specific assay. For the paper-based readout, a sample was considered positive when its test (T) band intensity was higher than that of the negative controls for that assay.

### Duplex SHINE reactions

The duplex IAV and RNase P SHINE assay was prepared with the following master mix: 1X SHINE buffer or LYO buffer, 45 nM *Lwa*Cas13a, 20nM *Lba*Cas12a, 62.5 nM 6rU quenched FAM reporter, 250nM 8C quenched HEX reporter, 2 mM of each rNTP, 1 unit/μL murine RNase inhibitor, 1 unit/μL NextGen T7 RNA polymerase, 90nM SARS-CoV-2 S gene RPA primers, 90nM RNase P RPA primers, 22.5 nM Cas12a crRNA specific against RNase P and 22.5 nM Cas13a crRNA specific against IAV. This master mix was used to resuspend the TwistAmp Basic Kit RPA pellets (1 pellet per 102 μL final master mix volume). Then, 0.1 unit/μL RNase H, 2 units/μL SuperScript IV reverse transcriptase and 14 mM magnesium acetate were added to the mastermix. For lyophilized SHINE pellets, magnesium acetate was omitted and the master mix was aliquoted into 65.5µL portions, flash frozen in liquid nitrogen and lyophilized overnight as described above. Duplex SHINE pellets were rehydrated prior to use. Contrived samples or patient samples were added to the SHINE reaction in a 1:19 ratio. Fluorescence intensity was measured on a Biotek Cytation 5 plate reader in two separate fluorescence channels (FAM: excitation: 485 nm, emission 520 nm; HEX: excitation 530 nm, emission 580 nm). For patient samples, a positive result was assigned to any sample with fluorescence values above the mean value plus 5 standard deviations for the negative (NTC) control.

The duplex oseltamivir resistance H275Y detection assay was carried out using the same protocol as above. Of note, both ancestral and H275Y strains share the same forward and reverse RPA primers so the master mix contained 90 nM of the shared RPA primer set. Both Cas13a crRNA targeting ancestral / Cas12a crRNA targeting H275Y and Cas13a crRNA targeting H275/ Cas12a targeting ancestral were tested in duplex SHINE format. Only the Cas13a crRNA targeting ancestral / Cas12a crRNA targeting H275Y was tested in lyophilized format and against contrived seedstock targets due to its better performance.

### Data analysis

Fluorescence values are reported as background-subtracted, with the fluorescence value measured before reaction progression (usually t = 5 min) subtracted from the final fluorescence value (90 min, unless otherwise indicated). Mean fluorescence values reported are the mean of 3 technical replicates, unless otherwise indicated. Data panels were generated using Python (version 3.7.4), seaborn (version 0.9.0) and matplotlib (version 3.1.1). The schematics shown in Figures 1a, 2a, 3a, 4a and Supplementary Figure 7 were created using www.biorender.com.

## Data Availability

All data produced in the present work are contained in the manuscript.

## Acknowledgements

We would like to thank members of the Myhrvold laboratory, Sabeti laboratory and the CDC’s Influenza Genomics lab - notably, C. Freije, H. Metsky, O. Dunkley, S. Siddiqui, O. Kimchi, L. Krasilnikova, T. Lan, J. Barnes and M. Kirby - for their valuable advice and reading of this manuscript; H. Metsky for his contributions to the assay design; J. Barnes and M. Kirby for kindly providing patient samples used in this study; and those researchers and laboratories who have made influenza sequencing data publicly available, which informed our assay design.

Funding was provided by the Defense Advanced Research Projects Agency (no. D18AC00006) and the Centers for Disease Control (no. 75D30122C15113). This work was also made possible by support from the Flu Laboratory and a cohort of generous donors through TED’s Audacious Project, including the ELMA Foundation, MacKenzie Scott, Skoll Foundation and Open Philanthropy. P.C.S. was also supported by the Merck KGaA Future Insight Prize.

## Author contributions

Y.B.Z. conceived the study under the guidance and supervision of P.C.S. and C.M. Y.B.Z. and T.- S.F.K.-T. performed initial experiments and data analysis to develop the two-step SHERLOCK assays for IAV and IBV. Y.B.Z. developed and optimized the one-step SHINE influenza assays. Y.B.Z., J.A.-S., and A.B performed and analyzed experiments with patient samples. J.A-S. provided critical insights on protocols, the results, and the work. Y.B.Z and J.A.-S. wrote the paper with guidance from P.C.S. and C.M. P.C.S and C.M. jointly supervised the project. All authors reviewed the manuscript.

