## Supplementary material for "CRISPR-based assays for point of need detection and subtyping of influenza"

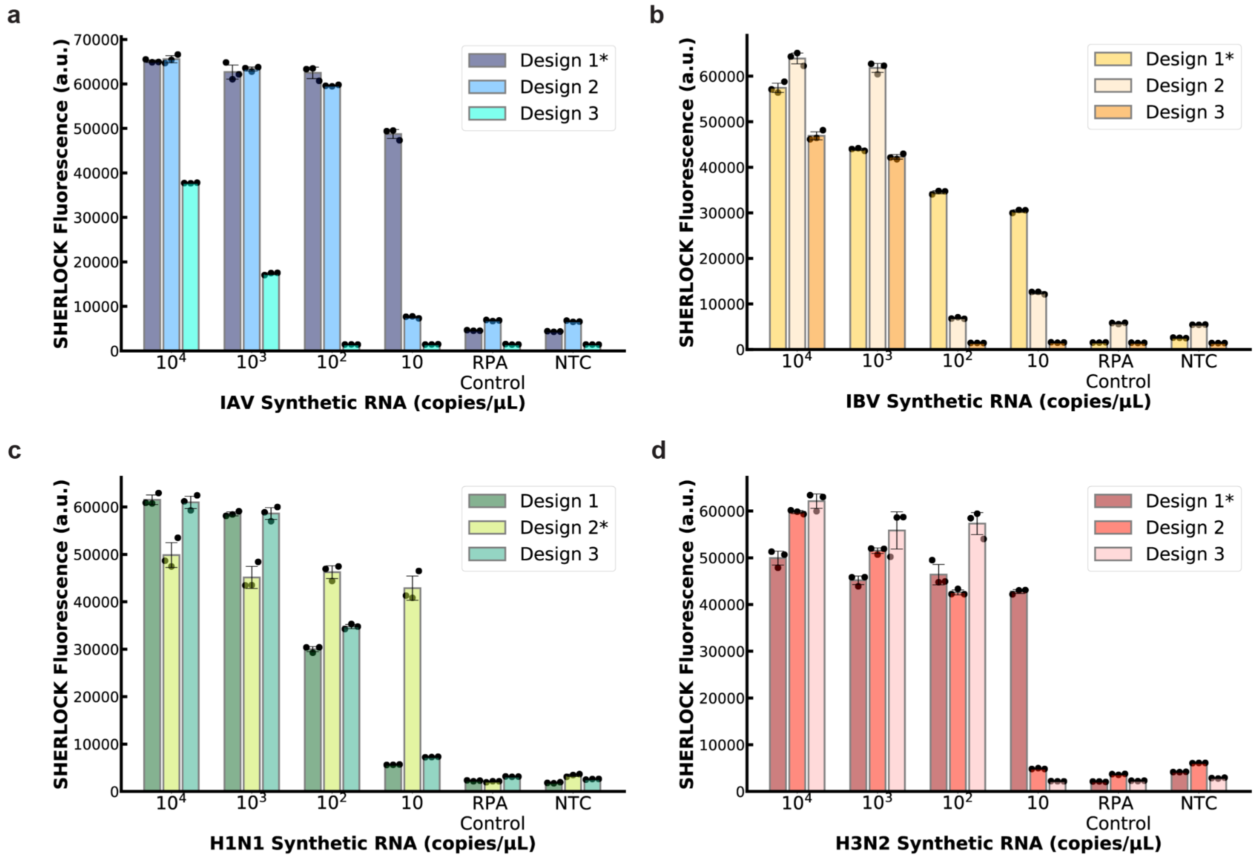


**Supplementary Figure 1: Testing the top three ADAPT designed primer-crRNA pairs for influenza assays with SHERLOCK.** SHERLOCK fluorescence of designs specific against **a)** IAV **b)** IBV **c)** H1N1 **d)** H3N2 synthetic RNA after 1.5 h at 37˚C. RPA control, nuclease free water input into RPA reaction, followed by standard SHERLOCK detection. NTC, no target control (nuclease free water input into SHERLOCK detection only without RPA). Values are mean fluorescence ± standard deviation of 3 technical replicates. * = design that was selected for further testing.

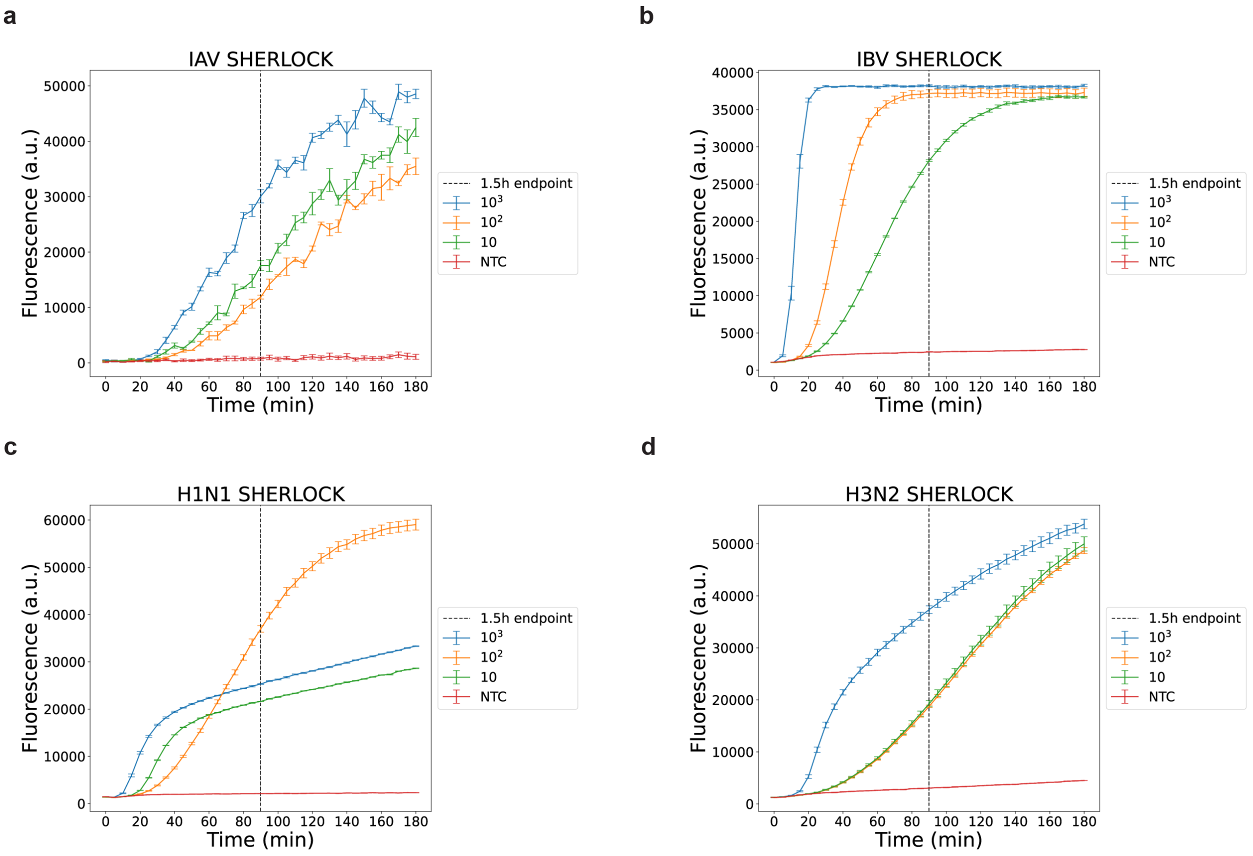


**Supplementary Figure 2: Fluorescence kinetics over time for the best performing SHERLOCK influenza assays. a)** IAV. **b)** IBV. **c)** H1N1. **d)** H3N2. Timepoints represent mean fluorescence ± standard deviation of 3 technical replicates.

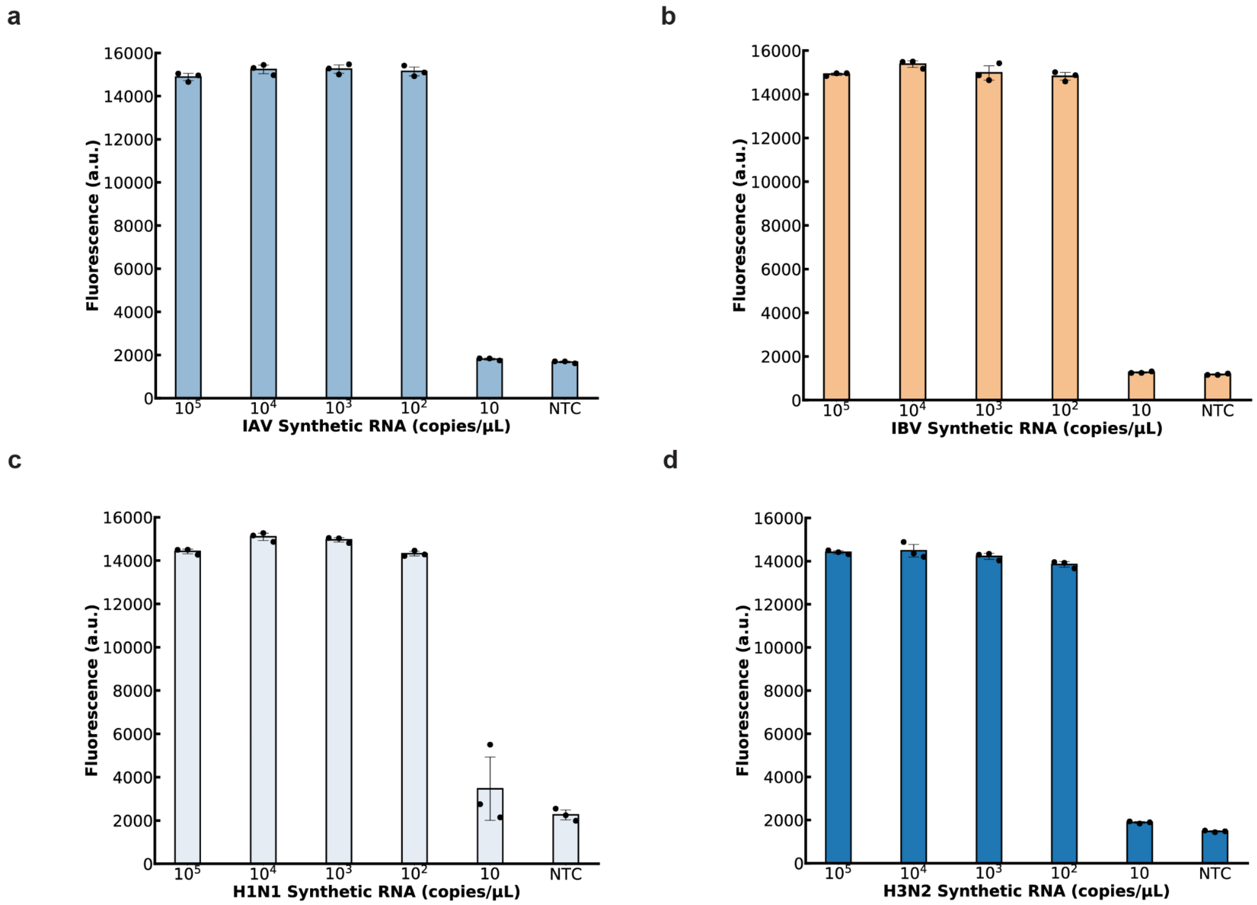


**Supplementary Figure 3: Developing single-reaction RPA amplification and Cas-13 detection assays for influenza.** Single-reaction fluorescent detection without lyophilization of **a)** IAV **b)** IBV **c)** H1N1 **d)** H3N2 synthetic RNA after 1.5 h at 37˚C. NTC, no target control. Values are mean fluorescence ± standard deviation of 3 technical replicates.

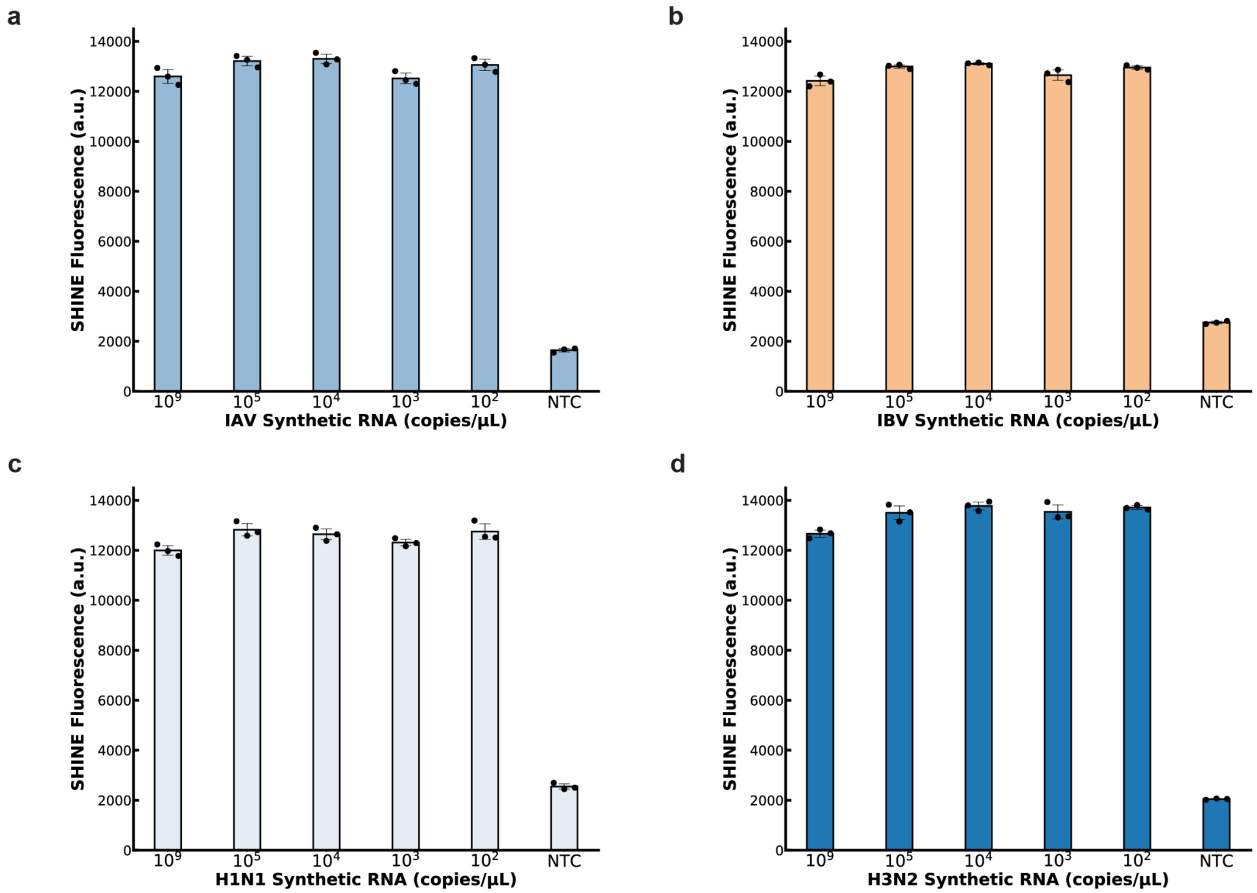


**Supplementary Figure 4: Developing influenza assays with the full SHINE platform workflow including lyophilization.** SHINE fluorescence of **a)** IAV **b)** IBV **c)** H1N1 **d)** H3N2 assays against their respective synthetic RNAs in UTM after 1.5 h at 37˚C. NTC, no target control. Values are mean fluorescence ± standard deviation of 3 technical replicates.

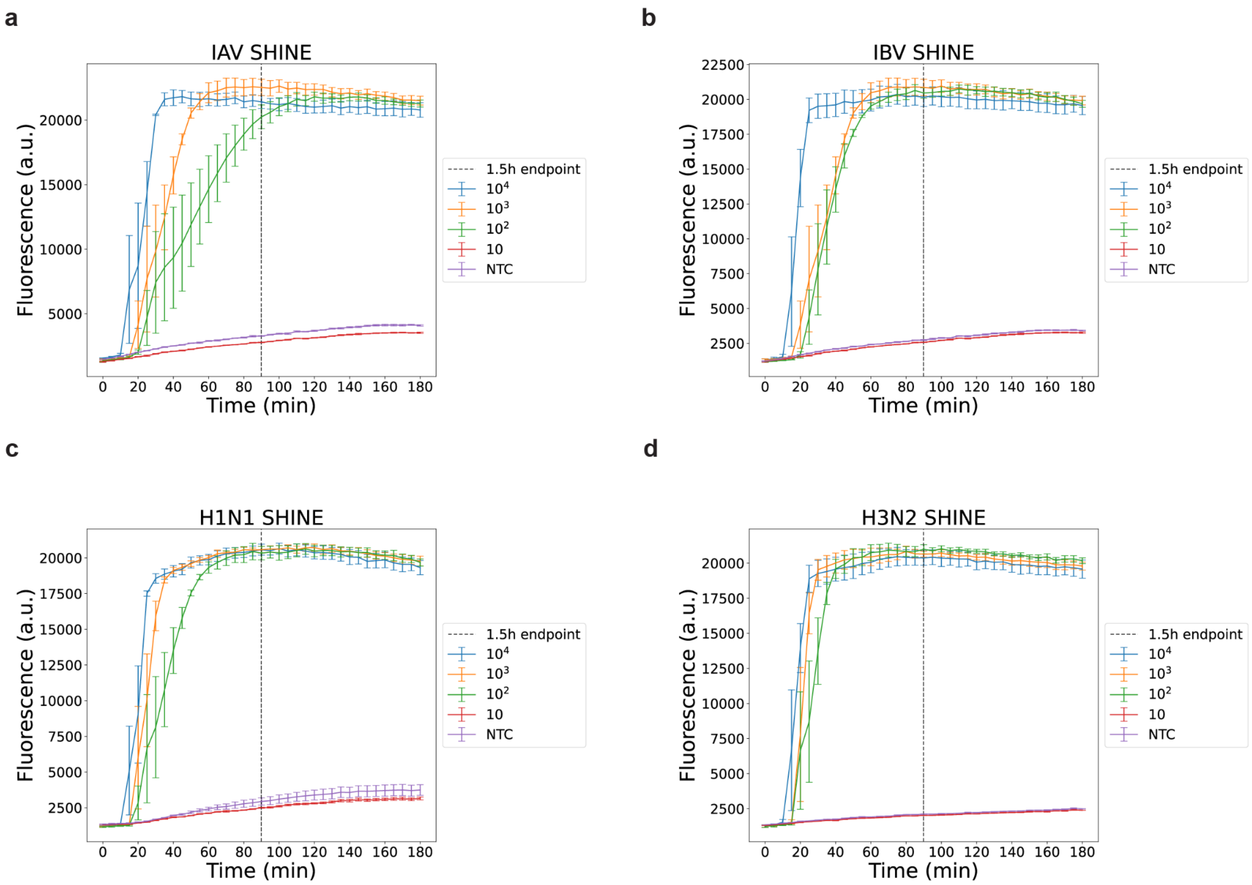


**Supplementary Figure 5: Fluorescence kinetics over time for SHINE influenza assays. a)** IAV. **b)** IBV. **c)** H1N1. **d)** H3N2. Timepoints represent mean fluorescence ± standard deviation of 3 technical replicates.

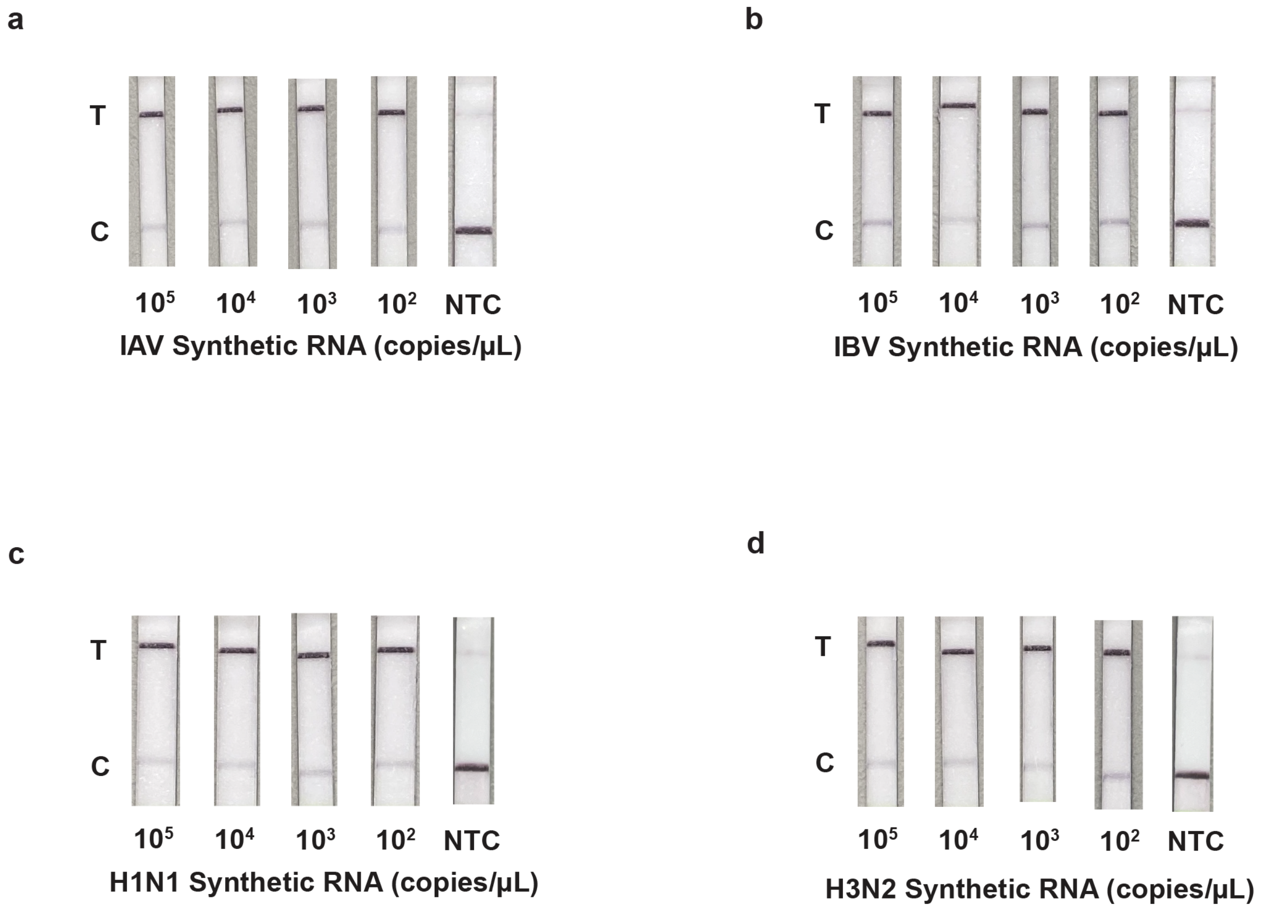


**Supplementary Figure 6: Influenza SHINE assays with paper-based readout.** SHINE detection with paper-based readout of **a)** IAV **b)** IBV **c)** H1N1 **d)** H3N2 assays against their respective synthetic RNAs after 1.5 h incubation at 37˚C. NTC, no target control. Shown are representative images of 3 technical replicates.

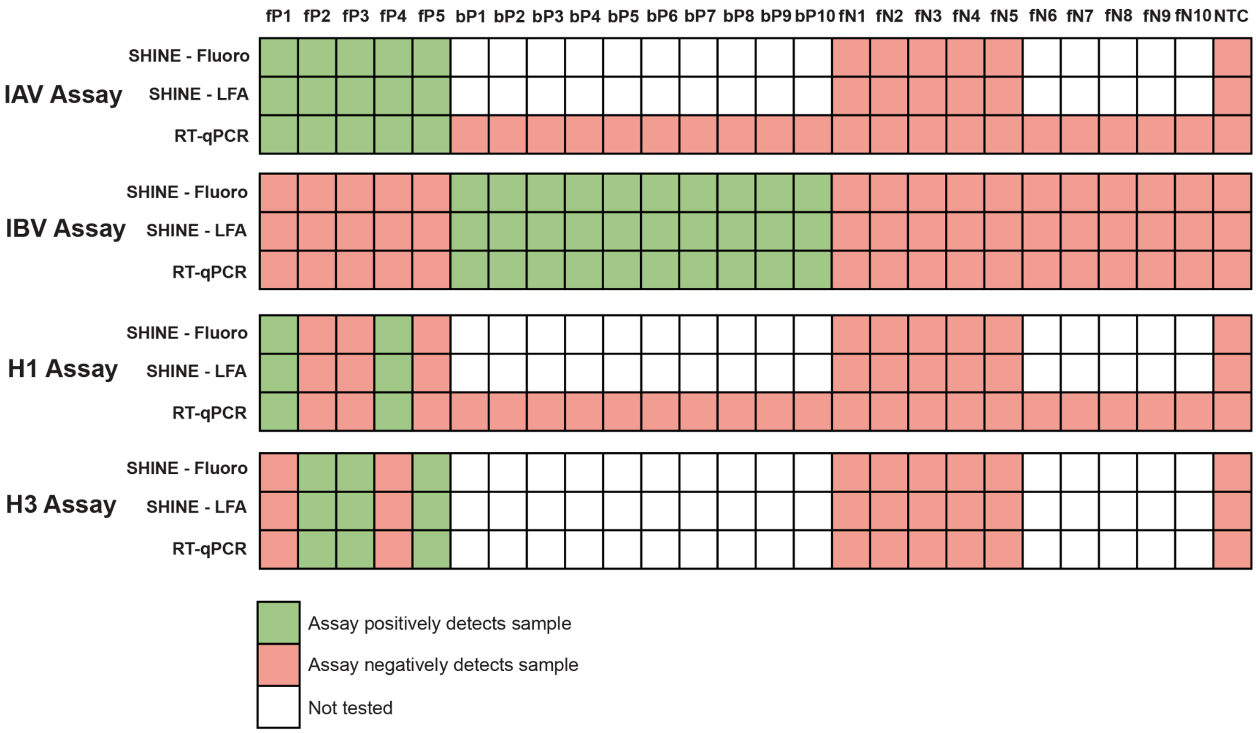


**Supplementary Figure 7: Summary of SHINE and RT-qPCR results on clinical samples.** . Fluoro: fluorescent readout. LFA: paper-based readout. NTC: no target control.

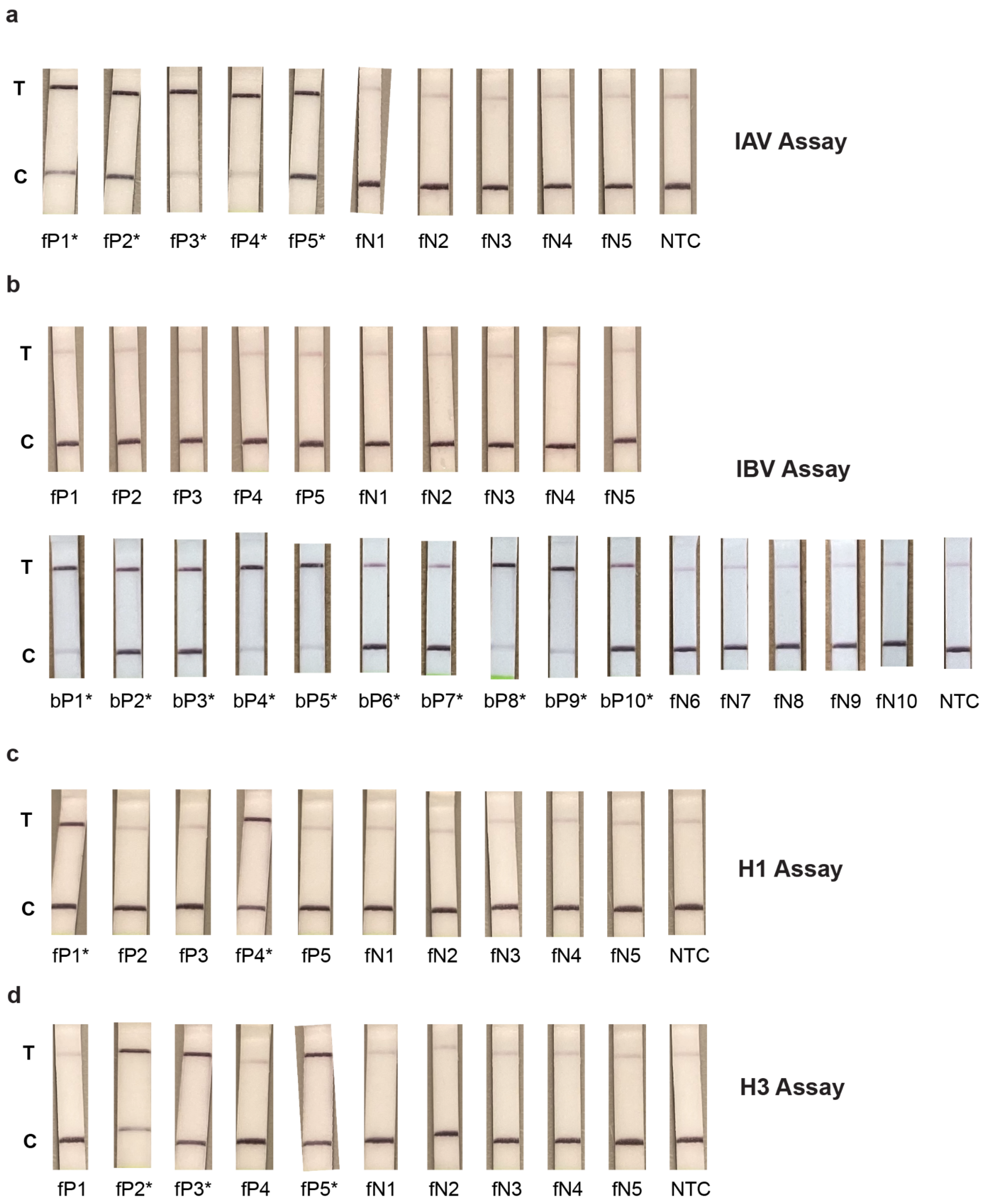


**Supplementary Figure 8: SHINE results on clinical samples with paper-based readout.** SHINE detection with paper-based readout of **a)** IAV **b)** IBV **c)** H1N1 **d)** H3N2 assays against clinical nasopharyngeal swabs after 1.5 h incubation at 37˚C. NTC, no target control. Shown are representative images of 3 technical replicates. * = qPCR positive sample.

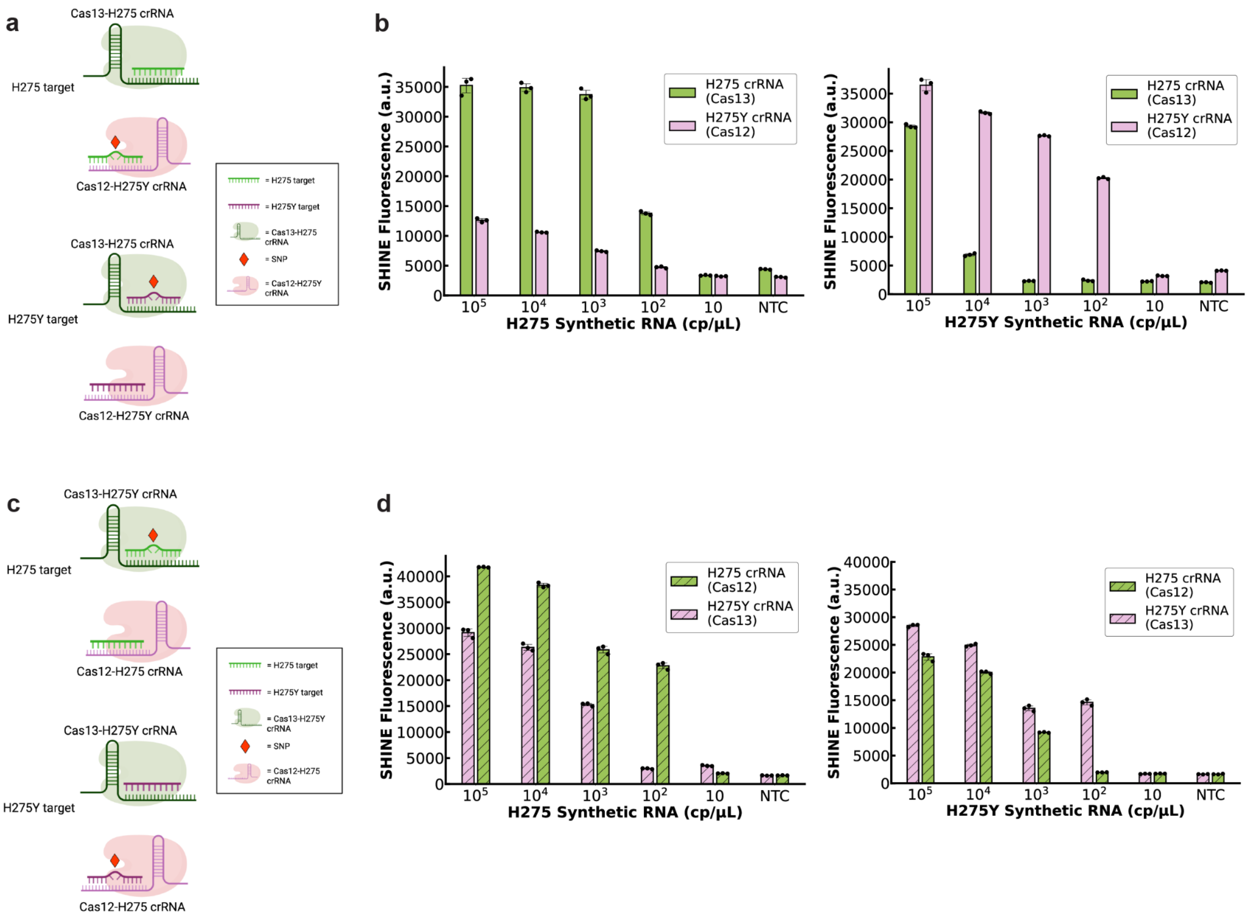


**Supplementary Figure 9: Testing the two possible configurations of CRISPR-Cas system for duplex SHINE detection. a,b)** Duplex SHINE assay with the Cas13 crRNA targeting the H275 oseltamivir-susceptible allele and the Cas12 crRNA targeting the H275Y oseltamivir-resistant allele. **a)** Schematic of crRNA-Cas pairings. **b)** This assay combination is tested on H275 and H275Y synthetic RNA targets. **c,d)** Duplex SHINE assay with the Cas13 crRNA targeting the H275Y oseltamivir-resistant allele and the Cas12 crRNA targeting the H275 oseltamivir-susceptible allele. **c)** Schematic of crRNA-Cas pairings. **d)** This assay combination is tested on H275 and H275Y synthetic RNA targets with fluorescent readout. Values are mean fluorescence ± standard deviation of 3 technical replicates after 1.5 h at 37˚C.

**Supplementary Table 1: Patient sample information.** CDC RT-qPCR validation of extracted patient samples. . RNase P is the assay internal control for extraction and detection quality control.. Ct scores shown as the mean of 3 technical replicates. n.d. = not detected. - indicates not tested. NTC: negative control

| Patient NP Swab | Flu A Ct score | Flu B Ct score | H1N1 Ct score | H3N2 Ct score | RNase P Ct score |
|---|---|---|---|---|---|
| fP1 | 23.085 | n.d. | 24.440 | n.d. | 24.232 |
| fP2 | 24.421 | n.d. | n.d. | 21.977 | 22.909 |
| fP3 | 21.712 | n.d. | n.d. | 24.385 | 23.548 |
| fP4 | 24.353 | n.d. | 25.343 | n.d. | 22.796 |
| fP5 | 25.540 | n.d. | n.d. | 26.830 | 23.175 |
| bP1 | n.d. | 24.296 | n.d. | - | - |
| bP2 | n.d. | 26.356 | n.d. | - | - |
| bP3 | n.d. | 26.712 | n.d. | - | - |
| bP4 | n.d. | 24.564 | n.d. | - | - |
| bP5 | n.d. | 23.495 | n.d. | - | - |
| bP6 | n.d. | 28.609 | n.d. | - | - |
| bP7 | n.d. | 31.085 | n.d. | - | - |
| bP8 | n.d. | 22.695 | n.d. | - | - |
| bP9 | n.d. | 24.121 | n.d. | - | - |
| bP10 | n.d. | 25.311 | n.d. | - | - |
| fN1 | n.d. | n.d. | n.d. | n.d. | 22.265 |
| fN2 | n.d. | n.d. | n.d. | n.d. | 22.810 |
| fN3 | n.d. | n.d. | n.d. | n.d. | 23.636 |
| fN4 | n.d. | n.d. | n.d. | n.d. | 23.049 |
| fN5 | n.d. | n.d. | n.d. | n.d. | 22.260 |
| fN6 | n.d. | n.d. | n.d. | - | - |

| | | | | | |
|---|---|---|---|---|---|
| fN7 | n.d. | n.d. | n.d. | - | - |
| fN8 | n.d. | n.d. | n.d. | - | - |
| fN9 | n.d. | n.d. | n.d. | - | - |
| fN10 | n.d. | n.d. | n.d. | - | - |
| NTC | n.d. | n.d. | n.d. | n.d. | n.d. |

**Supplementary Table 2: Oligonucleotides used in this study.**

| Name | Oligo Type | Sequence | Assay |
|---|---|---|---|
| fluA_design1* | crRNA | GAUUUAGACUACCCCAAAAACGAAGGGGACUAA AACUGGUCUUGUCUUUAGCCAUUCCAUGAGA | FLUV A |
| fluA_design2 | crRNA | GAUUUAGACUACCCCAAAAACGAAGGGGACUAA AACUGGUGUAUUUCUUUUGUCCAAGAUUCAG | FLUV A |
| fluA_design3 | crRNA | GAUUUAGACUACCCCAAAAACGAAGGGGACUAA AACCGAUCUCGGCUUUGAGGGGGCCUGAUGG | FLUV A |
| fluB_design1* | crRNA | GAUUUAGACUACCCCAAAAACGAAGGGGACUAA AACUCCAAUGUUUUUGAUGCCUAGUGCUGCU | FLUV B |
| fluB_design2 | crRNA | GAUUUAGACUACCCCAAAAACGAAGGGGACUAA AACGGUCAAAUUCUUUCCCACCGAACCAACA | FLUV B |
| fluB_design3 | crRNA | GAUUUAGACUACCCCAAAAACGAAGGGGACUAA AACAGUUCUGCUUUGCCUUCUCCAUCUUCUG | FLUV B |
| H1_design1 | crRNA | GAUUUAGACUACCCCAAAAACGAAGGGGACUAA AACCCAGGGAGACUACCAGUACCAAUGAACU | A(H1N1) |
| H1_design2* | crRNA | GAUUUAGACUACCCCAAAAACGAAGGGGACUAA AACAAUGAUAAUACCAGAUCCAGCAUUUCUU | A(H1N1) |
| H1_design3 | crRNA | GAUUUAGACUACCCCAAAAACGAAGGGGACUAA AACGAUGUACAUUCUGAAAUGGGAGGCUGGU | A(H1N1) |
| H3_design1* | crRNA | GAUUUAGACUACCCCAAAAACGAAGGGGACUAA AACACUGAGGGUCUCCCAAUAGAGCAUCUAU | A(H3N2) |
| H3_design2 | crRNA | GAUUUAGACUACCCCAAAAACGAAGGGGACUAA AACUGACUCCAGUCCAAUUGAAGCUUUCAUU | A(H3N2) |
| H3_design3 | crRNA | GAUUUAGACUACCCCAAAAACGAAGGGGACUAA AACUUCCCAUAUCCUCAGCAUUUUCCCUCAG | A(H3N2) |
| RP_g2 | crRNA | UAAUUUCUACUAAGUGUAGAUAGGCCCAGCUGG CCCGCUGC | RNase P |

| | | | |
|---|---|---|---|
| wt_cas13* | crRNA | GAUUUAGACUACCCCAAAAACGAAGGGGACUAAAACUAACAGGAGCAUUCCUCAUAAUGAUAAU | A(H1N1) oseltamivir resistance |
| mut_cas12* | crRNA | UAAUUUCUACUAAGUGUAGAUUUAUUAUGAGGAAUGCUCCU | A(H1N1) oseltamivir resistance |
| wt_cas12 | crRNA | UAAUUUCUACUAAGUGUAGAUUCAUUAUGAGGAAUGCUCCU | A(H1N1) oseltamivir resistance |
| mut_cas13 | crRNA | GAUUUAGACUACCCCAAAAACGAAGGGGACUAAAACUAACAGGAGCAUUCCUCAUAAUAAUAAU | A(H1N1) oseltamivir resistance |
| fluA_d1_fwd* | RPA primer | GAAATTAATACGACTCACTATAGGGCTCAAAGCCGAGATCGCGCAGAGACTTGAA | FLUV A |
| fluA_d2_fwd | RPA primer | GAAATTAATACGACTCACTATAGGGATGATGATGGGCATGTTCAACATGCTAAGT | FLUV A |
| fluA_d3_fwd | RPA primer | GAAATTAATACGACTCACTATAGGGTGGCTAAAGACAAGACCAATTCTGTCACCT | FLUV A |
| fluA_d1_rev* | RPA primer | CACGGTGAGCGTGAAAACAAACCCTAAAAT | FLUV A |
| fluA_d2_rev | RPA primer | TCTGAGGATTGGAGCCCATCCCACCAGTAT | FLUV A |
| fluA_d3_rev | RPA primer | GAGGCCCATGCAACTGGCAAGTGCACCAGC | FLUV A |
| fluB_d1_fwd* | RPA primer | GAAATTAATACGACTCACTATAGGGAGGCAAAGCAGAACTAGCAGAAAAATTACA | FLUV B |
| fluB_d2_fwd | RPA primer | GAAATTAATACGACTCACTATAGGGTGTTTAATATGCTATCTACCGTGTTGGGAG | FLUV B |
| fluB_d3_fwd | RPA primer | GAAATTAATACGACTCACTATAGGGTGGATAAAAAACAAAAGATGCTTAACTGAT | FLUV B |
| fluB_d1_rev* | RPA primer | TTTTAAAAAGCAGATAGAGGCACCAATTAG | FLUV B |

| | | | |
|---|---|---|---|
| fluB_d2_rev | RPA primer | TCAGAAGATTGCAGTCCATCCCATAAGTAT | FLUV B |
| fluB_d3_rev | RPA primer | GACCATGAGACAATATAGTAGCGCTGAGCT | FLUV B |
| h1_d1_fwd | RPA primer | GAAATTAATACGACTCACTATAGGGTCTACCAGATTTTGGCGATCTATTCAACTG | A(H1N1) |
| h1_d2_fwd* | RPA primer | GAAATTAATACGACTCACTATAGGGAGTGGTACCGAGATATGCATTCACAATGGA | A(H1N1) |
| h1_d3_fwd | RPA primer | GAAATTAATACGACTCACTATAGGGAACTTGTCAGACACCCGAGGGTGCTATAAA | A(H1N1) |
| h1_d1_rev | RPA primer | CATTAGAGCACATCCAGAAGCTGATTGCCC | A(H1N1) |
| h1_d2_rev* | RPA primer | TGTATTGCAATCGTGGACTGGTGTATCTGA | A(H1N1) |
| h1_d3_rev | RPA primer | TTGTGCTTTTTACATACTTTGGACATTTCC | A(H1N1) |
| h3_d1_fwd* | RPA primer | GAAATTAATACGACTCACTATAGGGCAGATCCTTGATGGAGAAAACTGCACACTA | A(H3N2) |
| h3_d2_fwd | RPA primer | GAAATTAATACGACTCACTATAGGGTTGCCTCATCCGGCACACTGGAGTTTAACA | A(H3N2) |
| h3_d3_fwd | RPA primer | GAAATTAATACGACTCACTATAGGGAACAAACTGTTTGAAAAAACAAAGAAGCAA | A(H3N2) |
| h3_d1_rev* | RPA primer | GGTCCCATTTCTTATTTTGAAAGCCATCAC | A(H3N2) |
| h3_d2_rev | RPA primer | CTTATGCAAGCAGAACTTGTTCCGTTTTGA | A(H3N2) |
| h3_d3_rev | RPA primer | TTGTGCTTTTTACATACTTTGGACATTTCC | A(H3N2) |
| RP_30nt_fwd | RPA primer | GTGGAATACACCCTTAGGAAAAGGCTTC | RNase P |
| RP_40nt_fwd* | RPA primer | GAACTGGATCCAGTGGAATACACCCTTAGGAAAAGGCTTC | RNase P |
| RP_50nt_fwd | RPA primer | GGATCCAGTGGAATACACCCTTAGGAAAAGGCTTCCCAGCCGCCTGCCCC | RNase P |

| | | | |
|---|---|---|---|
| RP_30nt_rev | RPA primer | CAAGCCGTGAATGTAGATCTCAGAGCAC | RNase P |
| RP_40nt_rev* | RPA primer | GGCCAGGCCCAAGCCGTGAATGTAGATCTCAGAGCA CGCG | RNase P |
| RP_50nt_rev | RPA primer | GGTTGATGGCCAGGCCCAAGCCGTGAATGTAGATCT CAGAGCACGCGTTC | RNase P |
| or_fwd | RPA primer | GAAATTAATACGACTCACTATAGGGTACAAAATCTT CAGAATAGAAAAGGGAAAG | A(H1N1) oseltamivir resistance |
| or_rev | RPA primer | CAGTTATCCCTGCACACACATGTGATTTCA | A(H1N1) oseltamivir resistance |
| or_fwd40nt | RPA primer | AGGCCTCATACAAAATCTTCAGAATAGAAAAGGGAA AGAT | A(H1N1) oseltamivir resistance |
| or_rev40nt | RPA primer | GCCAGTTATCCCTGCACACACATGTGATTTCACTAG AATC | A(H1N1) oseltamivir resistance |
| Flu A Synthetic RNA target 1* | gBlock | GAAATTAATACGACTCACTATAGGGGCCTTCTAACC GAGGTCGAAACGTATGTTCTCTCTATCGTTCCATCA GGCCCCCTCAAAGCCGAGATCGCGCAGAGACTTGAA GATGTCTTTGCTGGGAAAAACACAGATCTTGAGGCT CTCATGGAATGGCTAAAGACAAGACCAATTCTGTCA CCTTTGACTAAGGGGATTTTAGGGTTTGTTTTCACG CTCACCGTGCCCAGTGAGCGAGGACTGCAGCGTAGA CGCTTTGTCCAAAATGCCCTCAATGGGAATGGAGAC CCAAATAACATGGACAAAGCAGTTAAACTGTATAGG AAACTTAAGAGGGAGATAACGTTCCACGGGGCCAAA GAAATAGCTCTCAGTTATTCTGCTGGTGCACTTGCC AGTTGCATGGGCCTCATATACAATAGGATGGGGGCT GTAACCACTGAAGTGGCATTTGGCCTGGTGTGTGCA ACATGTGAGCAGA | FLUV A |
| Flu A Synthetic RNA target 2 | gBlock | GAAATTAATACGACTCACTATAGGGGTAAGAGAATG AAGCTCCGGACACAAATACCTGCAGAAATGCTAGCA AGCATTGACCTGAAGTATTTCAATGAATCAACAAGG AAGAAAATTGAGAAAATAAGGCCTCTTCTAATAGAT | FLUV A |

| | | | |
|---|---|---|---|
| | | GGCACAGCATCATTGAGCCCTGGAATGATGATGGGC<br>ATGTTCAACATGCTAAGTACAGTTTTAGGAGTCTCG<br>ATACTGAATCTTGGACAAAAGAAATACACCAAGACA<br>ACATACTGGTGGGATGGGCTCCAATCCTCAGACGAT<br>TTTGCCCTCATAGTGAATGCACCAAATCATGAGGGA<br>ATACAAGCAGGAGTGGATAGATTCTATAGGACCTGC<br>AAGTTAGTGGGAATCAACATGAGCAAAAAGAAGTCC<br>TATATAAATAAAACAGGGACATTTGAATTCACTAGC<br>TTTTTTTATCGATATGGATTTGTGGCTAATTTTAGC<br>ATGGAGCT | |
| Flu B Synthetic RNA target 1* | gBlock | GAAATTAATACGACTCACTATAGGGAAGGCTCAAAT<br>ACCTTGTCCTGATCTGTTCAGCATACCATTAGAAAG<br>ATATAATGAAGAAACAAGGGCGAAATTAAAAAGGCT<br>GAAGCCATTCTTCAATGAAGAAGGAACAGCATCTTT<br>GTCGCCTGGGATGATGATGGGAATGTTTAATATGCT<br>ATCTACCGTGTTGGGAGTAGCAGCACTAGGCATCAA<br>AAACATTGGAAACAAGGAATACTTATGGGATGGACT<br>GCAATCTTCTGATGATTTTGCTTTGTTTGTTAATGC<br>AAAAGATGAAGAAACATGTATGGAAGGGATAAATGA<br>TTTTTACCGAACATGTAAATTATTGGGAATAAACAT<br>GAGCAAAAAGAAAAGTTACTGTAACGAAACTGGAAT<br>GTTTGAATTTACAAGCATGTTCTATAGAGATGGATT<br>TGTATCTAACTTTGCAATGGAAATTCCTTCATTTGG<br>AGTTGCTG | FLUV B |
| Flu B Synthetic RNA target 2 | gBlock | GAAATTAATACGACTCACTATAGGGGCTGTTTGGAG<br>ACACAATTGCCTACCTGCTTTCATTGACAGAAGATG<br>GAGAAGGCAAAGCAGAACTAGCAGAAAAATTACACT<br>GTTGGTTCGGTGGGAAAGAATTTGACCTAGACTCTG<br>CCTTGGAATGGATAAAAAACAAAAGATGCTTAACTG<br>ATATACAGAAAGCACTAATTGGTGCCTCTATCTGCT<br>TTTTAAAACCCAAAGACCAGGAAAGAAAAAGAAGAT<br>TCATCACAGAGCCCTTATCGGGAATGGGAACAACAG<br>CAACAAAAAAGAAGGGCCTGATTCTGGCTGAGAGAA<br>AAATGAGAAAATGTGTGAGCTTTCATGAAGCATTTG<br>AAATAGCAGAAGGCCATGAAAGCTCAGCGCTACTAT<br>ATTGTCTCATGGTCATGTACCTGAATCCTGGAAATT<br>ATTCAATGCAAGTAAAACTAGGAACGCTCTGTGCTT<br>TGTGCGAAAAACA | FLUV B |
| H1 Synthetic RNA target 1 | gBlock | GAAATTAATACGACTCACTATAGGGATCTAAATAAA<br>AAAGTTGATGATGGTTTCCTGGACATTTGGACTTAC<br>AATGCCGAACTGTTGGTTCTACTGGAAAATGAAAGA<br>ACTTTGGACTATCACGATTCAAATGTGAAGAACTTG | A(H1N1) |

| | | | |
|---|---|---|---|
| | | TATGAAAAAGTAAGAAACCAGTTAAAAAACAATGCC AAGGAAATTGGAAACGGCTGCTTTGAATTTTACCAC AAATGCGATAACACATGCATGGAAAGTGTCAAGAAT GGGACTTATGACTACCCAAAATACTCAGAGGAAGCA AAATTAAACAGAGAAAAAATAGATGGAGTAAAGCTG GAATCAACAAGGATCTACCAGATTTTGGCGATCTAT TCAACTGTCGCCAGTTCATTGGTACTGGTAGTCTCC CTGGGGGCAATCAGCTTCTGGATGTGCTCTAATGGG TCTCTACAGTGTAGAATATGTATTTAACATTAGGAT TTCAGAA | |
| H1 Synthetic RNA target 2* | gBlock | GAAATTAATACGACTCACTATAGGGCAAGAAGGGAG AATGAACTATTACTGGACACTAGTAGAGCCGGGAGA CAAAATAACATTCGAAGCAACTGGAAATCTAGTGGT ACCGAGATATGCATTCACAATGGAAAGAAATGCTGG ATCTGGTATTATCATTTCAGATACACCAGTCCACGA TTGCAATACAACTTGTCAGACACCCGAGGGTGCTAT AAACACCAGCCTCCCATTTCAGAATGTACATCCGAT CACAATTGGGAAATGTCCAAAGTATGTAAAAAGCAC AAAATTGAGACTGGCCACAGGATTGAGGAATGTCCC GTCTATTCAATCTAGAGGCCTATTCGGGGCCATTGC CGGCTTCATTGAAGGGGGGTGGACAGGGATGGTAGA TGGATGGTACGGTTATCACCATCAAAATGAGCAGGG GTCAGGATATGCAGCCGATCTGAAGAGCACACAAAA TGCCATT | A(H1N1) |
| H3 Synthetic RNA target | gBlock | GAAATTAATACGACTCACTATAGGGGTTGGTTCAGA ATTCCTCAATAGGTGAAATATGCGACAGTCCTCATC AGATCCTTGATGGAGAAAACTGCACACTAATAGATG CTCTATTGGGAGACCCTCAGTGTGATGGCTTTCAAA ATAAGAAATGGGACCTTTTTGTTGAACGAAGCAAAG CCTACAGCAACTGTTACCCTTATGATGTGCCGGATT ATGCCTCCCTTAGGTCACTAGTTGCCTCATCCGGCA CACTGGAGTTTAACAATGAAAGCTTCAATTGGACTG GAGTCACTCAAAACGGAACAAGTTCTGCTTGCATAA GGAGATCTAGTAGTAGTTTCTTTAGTAGATTAAATT GGTTGACCCACTTAAACTACACATATCCAGCATTGA ACGTGACTATGCCAAACAATGAACAATTTGACAAAT TGTACATTTGGGGGGTTCACCACCCGGGTACGGACA AGGACCA | A(H3N2) |
| RP Synthetic RNA target | gBlock | GAAATTAATACGACTCACTATAGGGGTGCTGTG GAGGCTGAACTGGATCCAGTGGAATACACCCTT AGGAAAAGGCTTCCCAGCCGCCTGCCCCGGAGA | RNase P |

| | | | |
|---|---|---|---|
| | | CCCAATGACATTTATGTCAACATGAAGACGGAC<br>TTTAAGGCCCAGCTGGCCCGCTGCCAGAAGCTG<br>CTGGACGGAGGGGCCCGGGGTCAGAACGCGTGC<br>TCTGAGATCTACATTCACGGCTTGGGCCTGGCC<br>ATCAACCGCGCCATCAACATCGCGCTGCAGCTG<br>CAGGCGGGCAGCTTCGGGTCCTTGCAGGTGGCT<br>GCCAATACCTCCACCGTGGAGCTTGTTGATGAG<br>CTGGAGCCAGAGACCGACACACGGGAGCCACTG<br>ACTCGGATCCGCAACAACTCAGCCATCCACATC<br>CGAGTCTTCAGGGTCACACCCAAGTAATTGAAA<br>AGACACTCCTCCACTTATCCCCTCCGTGATATG<br>GCTCTTCGCATGCTGAGTA | |
| wt Synthetic RNA target | gBlock | GAAATTAATACGACTCACTATAGGGGTGATGGACAG<br>GCCTCATACAAAATCTTCAGAATAGAAAAGGGAAAG<br>ATAATCAAATCAGTCGAAATGAAAGCCCCTAATTAT<br>CACTATGAGGAATGCTCCTGTTACCCTGATTCTAGT<br>GAAATCACATGTGTGTGCAGGGATAACTGGCATGGC<br>TCGAATCGACCGTGGGTGTCTTTCAACCAGAATCTG<br>GAATATCAGATGGGATACATATGCAGTGGGGTTTTC<br>GGAGACAATCCACGCCCTAATGATAAGACAGGCAGT<br>TGTGGTCCAGTATCGTCTAATGGAGCAAATGGAGTA<br>AAAGGATTTTCATTCAAATACGGCAATGGTGTTTGG<br>ATAGGGAGAACTAAGAGCATTAGTTCAAGAAAAGGT<br>TTTGAGATGATTTGGGATCCGAATGGATGGACTGGG<br>ACTGACAATAAATTCTCAATAAAGCAAGATATCGTA<br>GGAATAA | A(H1N1) oseltamivir resistance |
| H275Y Synthetic RNA target | gBlock | GAAATTAATACGACTCACTATAGGGGGACCAAGTGA<br>TGGACAGGCCTCATACAAAATCTTCAGAATAGAAAA<br>GGGAAAGATAATCAAATCAGTCGAAATGAAAGCCCC<br>TAATTATTACTATGAGGAATGCTCCTGTTACCCTGA<br>TTCTAGTGAAATCACATGTGTGTGCAGGGATAACTG<br>GCATGGCTCGAATCGACCGTGGGTGTCTTTCAACCA<br>GAATCTGGAATATCAGATGGGATACATATGCAGTGG<br>GGTTTTCGGAGACAATCCACGCCCTAATGATAAGAC<br>AGGCAGTTGTGGTCCAGTATCGTCTAATGGAGCAAA<br>TGGAGTAAAAGGATTTTCATTCAAATACGGCAATGG<br>TGTTTGGATAGGGAGAACTAAGAGCATTAGTTCAAG<br>AAAAGGTTTTGAGATGATTTGGGATCCGAATGGATG<br>GACTGGGACTGACAATAAATTCTCAATAAAGCAAGA<br>TATCGTA | A(H1N1) oseltamivir resistance |

| | | | |
|---|---|---|---|
| 5C-HEX reporter | Fluorescence DNA reporter | 5' - /5HEX/CCCCC/3IABkFQ/-3' | - |
| 6U-FAM reporter | Fluorescence RNA reporter | 5'- /56-FAM/rUrUrUrUrUrU/3IABkFQ/-3' | - |
| FAMBio 5C LFA reporter | LFA DNA reporter | 5′-/5BiosG/CCCCCCCCCCCCCC/36-FAM/-3′ | - |
| FAMBio 14U LFA reporter | LFA RNA reporter | 5′-/5BiosG/rUrUrUrUrUrUrUrUrUrUrUrUrUrU/36-FAM/-3′ | - |

*denotes oligos that were selected for further testing in the later assays.
